# Uncertainty and error in SARS-CoV-2 epidemiological parameters inferred from population-level epidemic models

**DOI:** 10.1101/2022.07.01.22277134

**Authors:** Dominic G. Whittaker, Alejandra D. Herrera-Reyes, Maurice Hendrix, Markus R. Owen, Leah R. Band, Gary R. Mirams, Kirsty J. Bolton, Simon P. Preston

**Author notes:** joint first authors (Dominic G. Whittaker, Alejandra D. Herrera-Reyes).

## Abstract

During the SARS-CoV2 pandemic, epidemic models have been central to policy-making. Public health responses have been shaped by model-based projections and inferences, especially related to the impact of various non-pharmaceutical interventions. Accompanying this has been increased scrutiny over model performance, model assumptions, and the way that uncertainty is incorporated and presented. Here we consider a population-level model, focusing on how distributions representing host infectiousness and the infection-to-death times are modelled, and particularly on the impact of inferred epidemic characteristics if these distributions are misspecified. We introduce an *SIR*-type model with the infected population structured by ‘infected age’, i.e. the number of days since first being infected, a formulation that enables distributions to be incorporated that are consistent with clinical data. We show that inference based on simpler models without infected age, which implicitly misspecify these distributions, leads to substantial errors in inferred quantities relevant to policy-making, such as the reproduction number and the impact of interventions. We consider uncertainty quantification via a Bayesian approach, implementing this for both synthetic and real data focusing on UK data in the period 15 Feb–14 Jul 2020, and emphasising circumstances where it is misleading to neglect uncertainty.

## 1 Introduction

A simple Susceptible-Infected-Removed (‘SIR’) epidemic model, sometimes termed the ‘general epidemic model’ (Diekmann and Heesterbeek, 2000), is defined as follows. Suppose that *S*_*i*_ and *I*_*i*_ are the number of susceptible and infectious individuals, respectively, on day *i*; and that between day *i* and *i* + 1 for some rate constants *λ* and *θ, λS*_*i*_*I*_*i*_ of the susceptibles become infectious, and *θI*_*i*_ of the infectious individuals become ‘removed’ (by death or recovery, so that they are no longer susceptible nor infectious). Though very simple, this model embodies important epidemiological principles and exhibits reasonable dynamics, including exponential growth in the early stages and decline once the susceptible population is suitably depleted. The model also connects to key epidemiological parameters such as the basic reproduction number, R_0_, defined as expected number of cases directly generated by one case in a population where all individuals are susceptible, for this model equal to *λ/θ*. The *SIR* model remains widely used for prediction and inference for epidemics, including SARS-CoV-2 (Britton, 2020; Dehning et al., 2020).

One of this simple model’s limitations, however, is that it does not distinguish the infectious individuals by how long they have been infected, termed the *infected age*. Consequently, it assumes that the infectiousness of infected individuals is constant, and therefore that the time between individuals becoming infected and then either recovering or dying follows an exponential (or geometric, in discrete time) distribution. These assumptions are strongly in conflict with clinical literature (Ferretti et al., 2020; Ganyani et al., 2020; Harrison et al., 2020; Verity et al., 2020), as we discuss in the following section. The principal contributions of this paper are to introduce an epidemic model which structured by “infected age”, and to use this model as a basis to infer quantities important to policy-making. Similar models incorporating structure by infected age have been investigated by (Hart et al., 2020) and (Diekmann et al., 2021) in the the context of the “forward” problem, i.e., understanding the impact on model dynamics, in contrast to our focus here on the “inverse” problem, i.e., of understanding the impact on inference based on data: our goal is to explore the inferential consequences of ignoring the infected-age structure, and to highlight circumstances when doing so gives misleading conclusions.

Quantities of policy-making relevance and of interest to infer include: the timing and impact of behavioural changes, especially relating to non-pharmaceutical interventions (NPIs) such as ‘lockdown’ measures; the effect of NPIs on epidemiological characteristics such as the effective reproduction number, ℛ_i_ (defined later); the timing and size of the peak in the number of new daily infections; and the number of individuals that remain susceptible at the end of an epidemic wave.

In Section 2 we describe the data available on the epidemic time-course and on changes in mobility patterns relating to response to NPIs. We describe an infected-age-structured model, which is the central model in the paper, and some simpler models for comparison of the types sometimes used for inferring characteristics of an epidemic. The results in Section 3 include an application of the model to compare competing hypotheses for epidemic decline; a simulation study investigating the impact of using non-infected-age-structured models for inference when the data arise from an infected-age-structured model; and full Bayesian inference focusing on UK data for the SARS-CoV-2 outbreak in spring 2020, including for the quantities listed in the preceding paragraph. Section 4 contains a concluding discussion.

## 2 Data, Models and Methods

### 2.1 Data

#### 2.1.1 Deaths associated with SARS-CoV-2

We focus on country-level SARS-CoV-2 mortality data for England and Wales for the period Feb-July 2020. The total population, denoted *N* in the following models, is 59.1 million (ONS, 2020). Various time-course data are available for this period, including the number of confirmed cases by specimen date, though such data are influenced by rapid changes to testing capacity (DHSC, 2020) and strategy (Dunn et al., 2020) that make them challenging to connect to variables in an epidemic model. For this reason we focus exclusively on data for SARS-CoV-2-associated deaths collated by date of death for England and Wales by the UK Office for National Statistics. These deaths data count registered deaths where COVID-19 was mentioned on the death certificate (ONS, 2020), with the first death occurring on 8 Mar 2020. Although death data are available beyond July 2020, we restrict ourselves to considering inferences that may be made at the conclusion of the first epidemic wave. The deaths data are plotted later in results Figure 3.

#### 2.1.2 Mobility data

The UK government announced on 23 Mar 2020 that a lockdown would be imposed entailing severe restrictions including that that no one should leave their place of residence except for some specific reasons (UK Government, 2020a). Over the next 5 months this legislation was revised approximately monthly, sequentially expanding the permissible reasons to leave one’s place of residence, whilst introducing guidance on COVID-secure workplaces (Department for Business, Energy & Industrial Strategy and Department for Digital, Culture, Media & Sport, 2020) and mandating face coverings in some public spaces (UK Government, 2020b). Publicly available data indicate the extent to which public behaviour altered in this period. Google mobility data, for example, indicate a relative change with respect to a normal baseline, in activity in various categories across the UK (Google, 2020), and provide a proxy for changes in mixing intensity in England & Wales over this period. Figure 1A shows the mean of ‘workplace’ and ‘transit’ categories computed daily from the UK Google mobility data, which motivates later modelling of a change in population mixing intensity with a step function to account for the introduction of NPIs.

**Figure 1:**
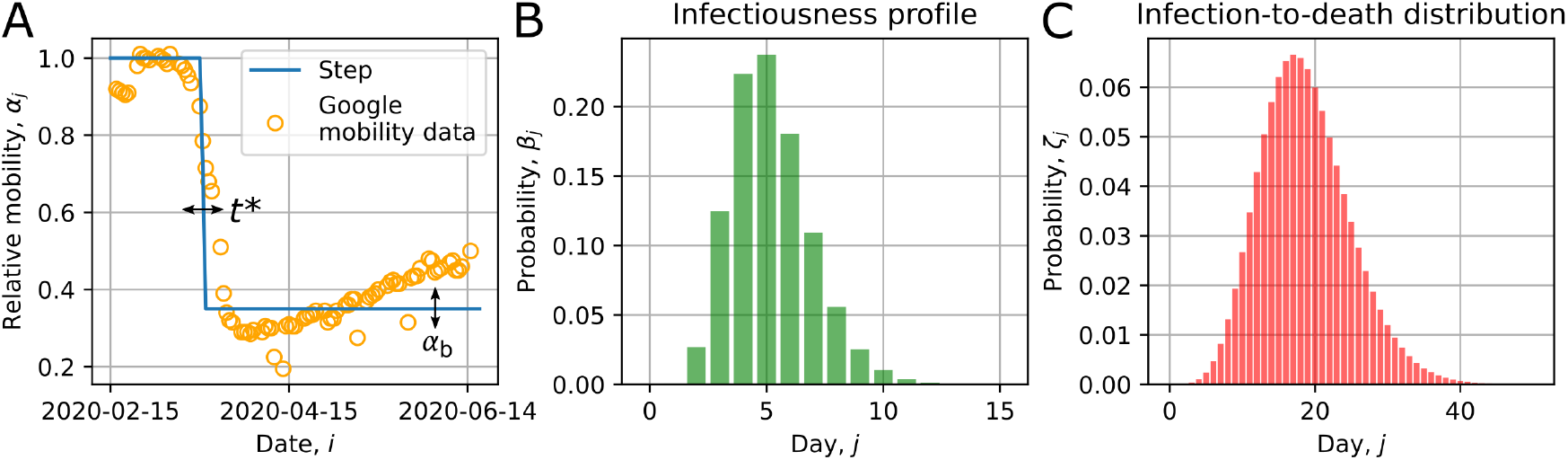
(A) Google mobility data and the parameterisation for a step function, defined in (3), for *α*_*j*_, which describes changes in population mixing intensity. The data points are given by the mean of workplace and transit activity levels for the UK, with Saturday and Sunday removed (the four outliers being bank holidays). The blue step is given by equation (3) and contains two parameters which control the timing and severity of the drop in viral transmission due to lockdown measures: *t*^∗^ and *α*_*b*_, respectively. Plots (B) and (C) respectively show the infectiousness profile, over a 15 day period, and the infection-to-death distribution, shown over a 50 day period; these correspond to the distributions and default parameters listed in Table 1.

#### 2.1.3 Clinical data on individuals’ response to SARS-CoV-2 infection

The models in §2.2 involve epidemiological parameters drawn from the clinical literature that characterise individuals’ infectiousness and mortality. These are summarised in Table 1, including ‘default’ values that are used in the later simulation study, and priors that are used for the Bayesian inference.

**Table 1:** Summary of key epidemiological parameters drawn from clinical literature, as described in §2.1.3, including default values used later to generate synthetic data (§3.2), and priors used in the Bayesian inference (§3.4). The default values correspond with the plots of the infectiousness profile and infection-to-death distribution shown in Figures 1B and C. The gamma distribution priors are parameterised as Γ(scale, shape), and the normal priors as 𝒩 (mean, std dev).

| Quantity | Distn | Param | Symbol | Default | Prior |
| --- | --- | --- | --- | --- | --- |
| Infectiousness profile, $\{\beta_j\}$ | $\Gamma$ | mean | $\beta_\mu$ | 5.2 | $\Gamma(16, 1/3)$ |
| | | variance | $\beta_{\sigma^2}$ | 2.96 | $\Gamma(2, 2.2)$ |
| Infection-to-death distribution, $\{p_j^\zeta\}$ | NB | mean | $\zeta_\mu$ | 18.69 | $\mathcal{N}(20, 3)$ |
| | | dispersion | $\zeta_\phi$ | 0.0546 | $\mathcal{N}(0.03, 0.025)$ |
| Infection fatality rate | — | — | $\delta$ | 0.00724 | $\mathcal{N}(0.00724, 0.001)$ |

For most hosts infected with SARS-CoV-2, detectable viral load in the upper respiratory tract peaks at around 5–6 days following exposure (He et al., 2020). For many hosts this approximately aligns with the the delay from exposure to experiencing symptoms (the *infection-to-onset* period) (Backer et al., 2020), and thus there is a significant period of infectiousness prior to symptom onset (He et al., 2020; Ashcroft et al., 2020). Some hosts remain asymptomatic throughout infection and experience similar viral load dynamics, besides somewhat faster viral clearance following peak viral load (Kissler et al., 2020). For the purposes of developing a transmission model, it is therefore important to describe a typical host *infectiousness profile, β*_*j*_, subject to Σ_*j*_ *β*_*j*_ = 1, as depending on the time in days, *j*, since infection (rather than with respect to symptom onset).

The generation time interval, defined as the period from host infection to the generation of progeny infection, has a distribution that describes such an infectiousness profile (Lehtinen et al., 2020). We use the inferred gamma-distributed generation time from the Singapore cluster in Ganyani et al. (2020), with mean 5.2 days and variance 2.96 days, to define a fixed infectiousness profile *β*_*j*_ when inferring the impact of lockdown in §3.4. Since the models in this paper are in discrete time, we discretise the gamma distribution by taking *β*_1_ = *F* (3*/*2) and *β*_*j*_ = *F* (*j* + 1*/*2) −*F* (*j* −1*/*2) for *j* ≥ 2, where *F* (·) is the gamma cumulative distribution function. We do not have generation time interval estimates for transmission chains specific to England and Wales. However estimates for other international data sets are congruent with the estimate from Ganyani et al. (2020). Ferretti et al. (2020) estimates a mean generation time of 5.0 days and variance 1.9 from a sample of 40 infector-infectee pairs. Sun et al. (2020) estimate a similar median generation time (5.3 days, IQR:3.1-8.7) that is correlated with the delay from symptom onset to isolation. To account for potential variability due to alternate estimates for the latent period, the role of an infected’s behaviour — in particular the efficacy and timing of isolation — in influencing the generation interval, and the potential for unrepresentative sampling of infector-infectee pairs, we draw on the range and reported confidence across these estimates to construct priors on the mean and variance of a gamma distributed infectiousness profile (see Table 1).

We refer to *β*_*j*_ as the infectiousness profile rather than the generation time distribution, as the latter has a small correction due to any overlap in the distributions for time from infection to death and infectiousness by infected-age due to the non-zero hazard of death while infectious (see §2.2.5). Figure 1B shows the ‘default’ infectiousness profile.

To infer the epidemic dynamics based on the death data, it is key to characterise accurately the *infection-to-death time distribution*. We denote the probability mass function of the infection-to-death distribution by 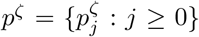, such that 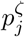 is the probability that death occurs *j* days after infection, conditional on ultimately dying from SARS-CoV-2. The infection-to-death distribution is the convolution of infection-to-onset and onset-to-death distributions, for which separately there are data.

The infection-to-onset period is estimated from cases with well constrained exposure history (e.g. Backer et al., 2020). For consistency with our choice of infectiousness profile, we adopt the infection-to-onset period used by Ganyani et al. (2020) when inferring the generation interval distribution above, namely a gamma distribution with mean 5.2 days and standard deviation 2.8 days. For onset to death, early data from Hubei, China, corrected for epidemic growth, suggested an average time 18 days and standard deviation 8.4 days (Verity et al., 2020). A report published by the UK government provides distributions for the time from symptom onset to death for over 22,000 patients hospitalised in the UK during the first epidemic wave (up August 1st 2020), by age range and sex (Harrison et al., 2020). The aggregate distribution is described by a gamma distribution with mean 14 days (std. dev. 9.8 days), and is stable when re-weighting the age- and sex-dependent distributions to match those of reported deaths in England and Wales over the same period (ONS, 2020), giving some confidence that this distribution is representative for the population we are modelling over the period of study. Similar estimates for the death delay are reported by Sherratt et al. (2021) based on confidential UK data.

We model the infection-to-death distribution as a negative binomial distribution, chosen for an appropriate shape, computing point estimates for its parameters by matching moments to the convolution of the Ganyani et al. (2020) infection-to-onset and Harrison et al. (2020) onset-to-death distributions described above. This leads to the parameters shown in Table 1 and the distribution plotted in Figure 1C. There remains uncertainty, however, in the infection-to-death distribution owing to uncertainty in the infection-to-onset period (see Backer et al., 2020), the censoring effect of unknown symptom onset dates in the hospitalisation data (Harrison et al., 2020), and regional variability (about which we lack data) that may influence the effective average time to death profiles (e.g. Hawryluk et al., 2020). We hence characterise uncertainty in the infection-to-death distribution via priors on the mean and dispersion parameters. We choose priors, shown in Table 1, such that infection-to-death distributions with high prior probability are consistent with the distributions estimated by Harrison et al. (2020) and Verity et al. (2020) time-to-death distributions.

A further key parameter for inferring epidemic dynamics from death data is the *infection fatality rate* (IFR), *δ*, which is the risk of death for an infected host, neglecting other host covariates. Seroprevalence of antibody to SARS-CoV-2 — together with evidence regarding the preservation of measurable antibody (Huang and Garcia-Carreras, 2020) — provides an estimate of the integrated exposure history to SARS-Cov-2, and enables estimation of the *δ*. In the results sections we explore the impact of different assumptions about *δ*: in §3.1 we let *δ* be a free parameter; in §3.2 we fix its value to the central estimate for England from (O’Driscoll et al., 2020); and then in §3.3 and §3.4 we try to characterise knowledge about *δ* with a judicious prior (Table 1) that reflects uncertainty and bias that may arise for various reasons, such as non-representative serological surveys, non-uniform prevalence in different risk groups (e.g. in care homes versus the surrounding community) and waning antibody levels (O’Driscoll et al., 2020).

### 2.2 Epidemic models

In the following we present several simple epidemic models. The first, called *SI*_*t*_*D*, is the central one in this paper and introduced with the goal of being as simple as possible whilst retaining structure by infected age. The subsequent *SIRD*_Δ*D*_, *SIURD* and *SE*^2^*I*^2^*U* ^2^*RD* models are simple models that are not directly structured by infected age. Figure 2 shows schematics of the various models.

**Figure 2:**
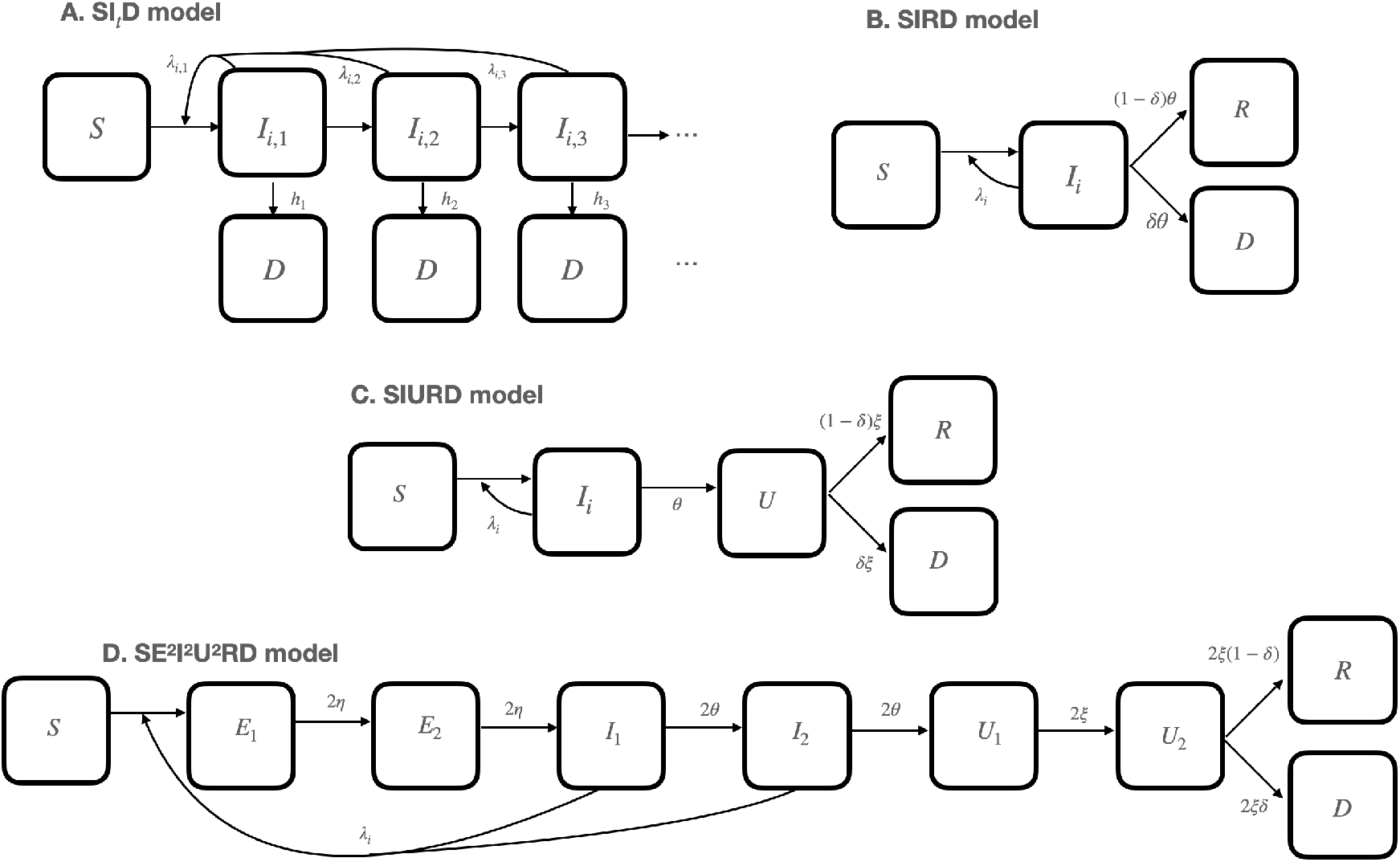
Compartmental structure for models (A) SI_t_D, (B) *SIRD*/*SIRD*_Δ*D*_ model, (C) *SIURD*, and (D) *SE*^2^*I*^2^*U* ^2^*RD*, as defined in §2.2. In (B) the *D*_Δ*D*_ compartment indicates deaths that will occur after an additional fixed delay Δ*D*. The *SIRD* model is the special case of the *SIRD*_Δ*D*_ model with Δ*D* = 0. Model (D) includes includes two compartments for each of the exposed (*E*_1_, *E*_2_), infectious (*I*_1_, *I*_2_), and non-infectious (*U*_1_, *U*_2_) states.

**Figure 3:**
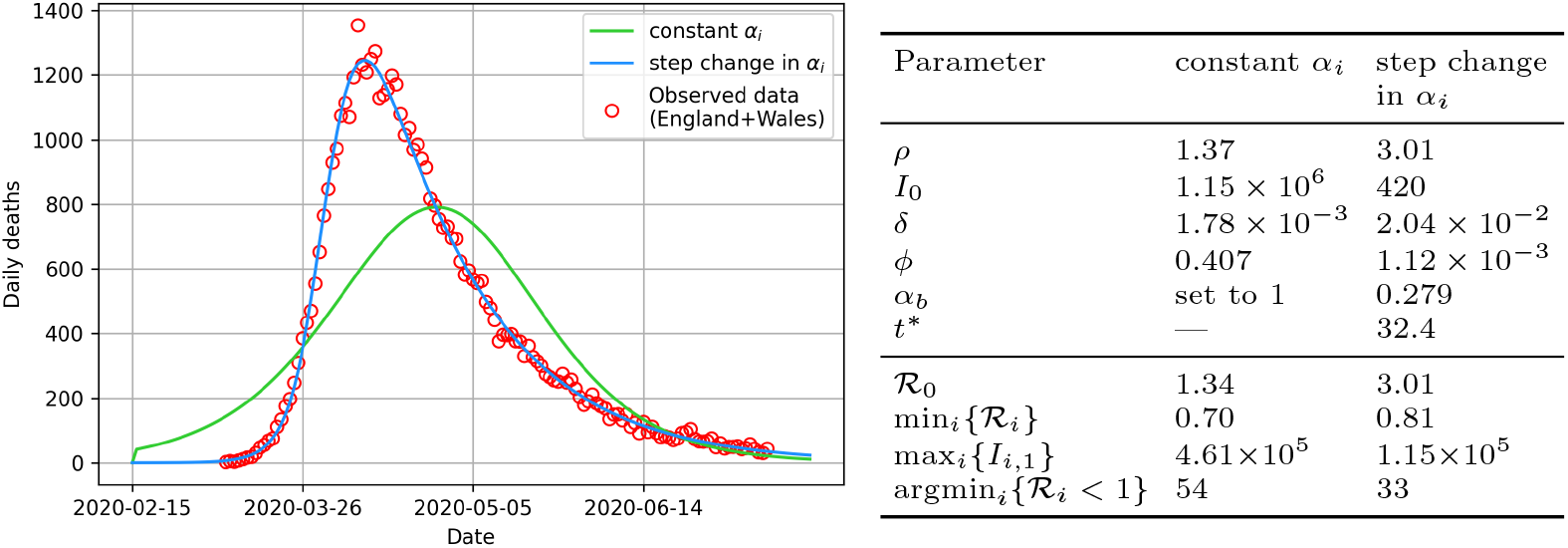
A comparison of the maximum likelihood parameter set SI_t_D model predictions for cases in which *ρ, I*_0_, the infection fatality rate, *δ*, and NB *ϕ* are inferred and infectivity is constant (herd immunity), which is a special case of the model in which step function parameters *t*^∗^ and *α*_*b*_ are also inferred (2 extra parameters; non-herd immunity). Inferred parameters are shown in the table to the right, together with emerging epidemiological quantities of interest below. The argmin_*i*_{ℛ_*i*_ *<* 1} values of 54 vs. 33 translate to 9th April vs. 19th March 2020, respectively.

#### 2.2.1 A model structured by infected age: *SI*_*t*_*D*

We denote by *I*_*i,j*_ the number of individuals who on day *i* became infected *j >* 0 days ago; and the total number of infected individuals on day *i* is thus *I*_*i*_ = Σ_*j*_ *I*_*i,j*_. In this model, between day *i* and *i* + 1 the number of new infections is *I*_*i*+1,1_ = *S*_*i*_Σ_*j*_ *λ*_*ij*_*I*_*i,j*_, for suitable *λ*_*ij*_ that in general depends on time *i* (to reflect changing transmission owing to changes in population mixing intensity, e.g., due to NPIs) and infected age *j* (to reflect non-constant infectiousness of those infected); and the number of new deaths amongst individuals with infected age *j* is *h*_*j*_*I*_*i,j*_, for suitable *h*_*j*_. The general infected-age-structured model is therefore

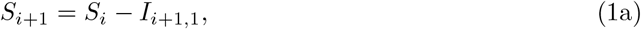

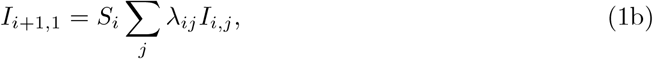

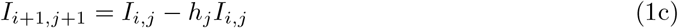

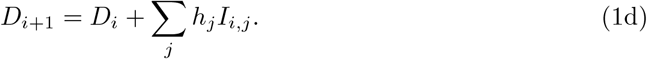

The degrees of freedom in the model need to be controlled via some further modelling choices for the {*λ*_*ij*_} and {*h*_*j*_}. We will write

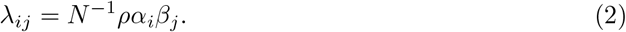

Here, *α*_*i*_ models the population mixing intensity relative to pre-epidemic social behaviour and is subject to the constraint *α*_1_ = 1. In practice *α* is a composite parameter capturing contact rates, social distancing (including mask wearing) and mobility. A simple model for the impact of introducing NPIs is to enable a sharp reduction in *α*_*i*_ at some change-point time *t*^∗^. Enabling a continuous *t*^∗^ helps in Markov Chain Monte Carlo procedures described later, hence we model *α*_*i*_ to transition from 1 to a new baseline *α*_*b*_ at change-point time *t*^∗^ via

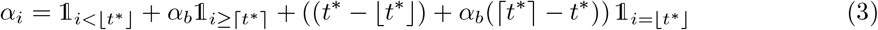

in which ⌊·⌋, ⌈·⌉ and 𝟙_(·)_ respectively denote the standard floor, ceiling and indicator functions. An example of {*α*_*i*_} is shown in Figure 1A.

The infectiousness profile *β*_*j*_, defined in §2.1.3, is a probability mass function where Σ_*j*_ *β*_*j*_ = 1, so in view of the constraints on {*α*_*i*_} and {*β*_*j*_}, the *ρ >* 0 is included in (2) as an intensity

parameter. Parameter *N* = *S*_*i*_ + *I*_*i*_ + *D*_*i*_ is the population size, which in this model is constant, because births and non-SARS-CoV-2 death are negligible on the time scales of interest and are thus neglected.

In the language of survival analysis, the {*h*_*j*_ : *j* ≥ 1} is the *hazard rate function* for the death of infecteds, such that *h*_*j*_ is the probability that an individual having survived *j* − 1 days post-infection will die on day *j*. In terms of the infection-to-death distribution, *p*_*ζ*_, and its corresponding cumulative distribution function 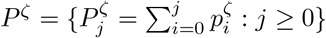,

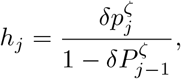

where *δ* is the infection fatality rate.

In this model, in contrast to some common epidemic models, including the non-infected-age-structured ones described below, the *I* denotes individuals who have been “infected” but are not necessarily “infectious”, since individuals of infected age *j* contribute to infecting susceptibles if and only if *β*_*j*_ *>* 0. This removes the need for an *E* variable for “exposed but pre-infectious” individuals, or an *R* variable for “recovered” individuals.

#### 2.2.2 *SIRD* and *SIRD*_Δ*D*_ models

A simple model and commonly used model (e.g. Britton, 2020; Lourençco et al., 2020) that does not maintain structure by infected age is as follows.

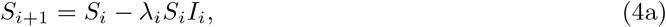

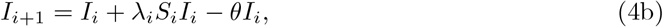

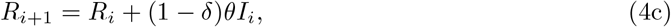

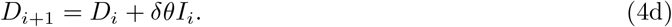

In this model (and those below), the number of new daily infecteds is *I*_*i*+1,1_ = *λ*_*i*_*S*_*i*_*I*_*i*_, where

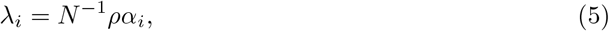

and *N* = *S*_*i*_ + *I*_*i*_ + *R*_*i*_ + *D*_*i*_. There is a close parallel to (1) but with key differences: infectiousness of infected individuals is constant with respect to infected age (*β*_*j*_ is taken to equal 1), and the hazard of removal from being infected is also constant (*h*_*j*_ is taken to equal some constant *θ*). This model includes an *R* variable, because the assumption of constant infectiousness of *I* individuals necessitates a way other than death for an infected individual to cease being infectious. By analogy to (1), the constant hazard implies that the duration in infected state is geometrically distributed.

The *SIRD*_Δ*D*_ model is a generalisation of the *SIRD* model that replaces equation (4d) with

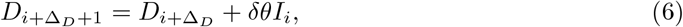

which introduces a non-negative integer “delay-to-death” parameter, Δ_*D*_. Introducing a fixed delay in this way is a common modelling strategy to make infection-to-recovery and infection-to-death distributions distinct (e.g. Lourencço et al., 2020) and the latter non-exponential. The basic *SIRD* model is the special case with delay Δ_*D*_ = 0. A generalisation of (6), in which Δ_*D*_ is not necessarily an integer, is

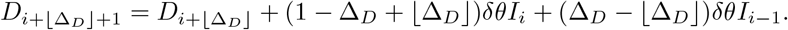

For example, if Δ_*D*_ = 7.2 then 80% of the deaths will have a delay of 7 days and 20% will have a delay of 8 days.

#### 2.2.3 An *SIURD* model

A different strategy besides incorporating a delay is to incorporate additional model states (see e.g. Royal Society SET-C (2020)). A model variation in this spirit is to incorporate a non-infectious state *U* that follows infection but precedes recovery or death, i.e.,

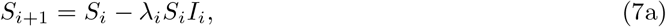

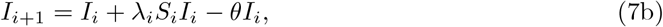

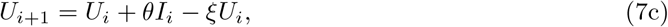

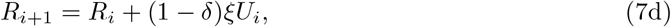

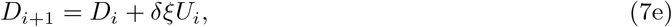

in which *ξ* is a rate constant.

#### 2.2.4 An *SE*^2^*I*^2^*U* ^2^*RD* model

The same strategy can be extended by including more states, for example an exposed (infected but not yet infectious) state, and by representing states using multiple compartments. Such an approach can enable the model dynamics to mimic the delay to peak infectiousness, and the delay between infection and death, and hence indirectly models infected ages (Hurtado and Kirosingh, 2019). The following model, chosen because it closely matches the approach of Kucharski et al. (2020), uses two compartments for each of the E, I and U states.

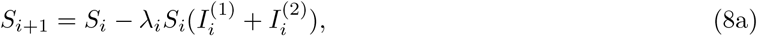

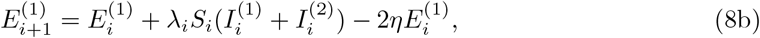

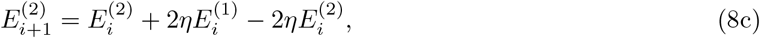

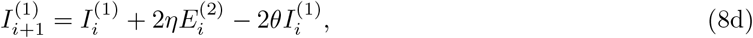

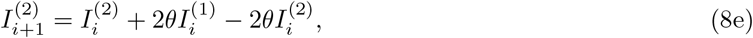

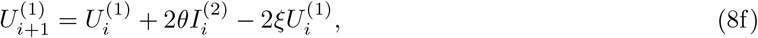

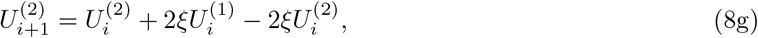

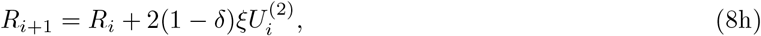

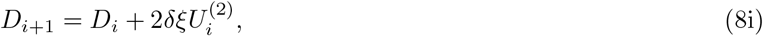

in which *η* is a rate constant.

#### 2.2.5 Connection to epidemiological parameters

The foregoing models connect directly to some key epidemiological parameters. A parameter important for characterising whether the epidemic is growing or declining is the time-varying reproduction number, ℛ_*i*_, of which there are multiple variations. We adopt for ℛ_*i*_ in this paper the instantaneous reproduction number (Fraser, 2007), which is the average number of secondary infecteds generated by a single infected assuming there are no population level changes in susceptibility or mixing behaviour during their infection. We calculate ℛ_*i*_ by performing the sum over infected age *j* of the number of new infecteds generated by a host at infected age *j* (Heffernan et al., 2005). For the *SI*_*t*_*D* model:

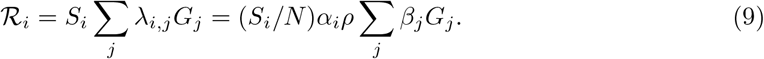

where *G*_*j*_ is the survival function of an infected individual associated with the hazard *h*_*j*_ of removal from the *I*, defined as the probability of not having been removed by day *j* after infection, which equals 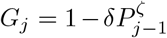. The product *β*_*j*_*G*_*j*_ is the proportion of the ℛ_*i*_ secondary infections that are generated at an infected age *j*, and therefore when normalised is the (discretised) generation time distribution.

For the *SIRD, SIRD*_Δ*D*_, *SIURD* and *SE*^2^*I*^2^*U* ^2^*RD* models, we compute ℛ_*i*_ from the same definition, noting that infecteds must transit through *I* before reaching *D* (*G*_*j*_ = 1). Given residence in an infectious state, the number of new infecteds generated per infected (*λ*_*i*_*S*_*i*_) is independent of infected-age. The infectiousness profile is *β*_*j*_ = *A*_*j*_*/* Σ (*A*_*j*_), in which *A*_*j*_ is the proportion of infecteds in an infectious state at infected-age *j*, which can be derived by considering the disease progression defined in equations (4), (7) or (8) as a Markov process (A). Hence *β*_*j*_ depends on the number of *E* and *I* states as well as the parameters governing the total time in the latent (*η*) and infected classes (*θ*). However, by construction, the average residence time in an infectious state is *θ*^−1^ for each of these models. Hence

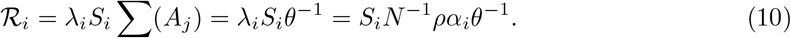

Estimates of ℛ_*i*_ can be used to infer the benefit of mitigating measures such as NPIs for reducing transmission. Of strategic interest is to infer argmin_*i*_ {ℛ_*i*_ *<* 1}, as this indicates the time *i* when NPIs were sufficient to put the epidemic into decline. Also related and important is the peak daily incidence, max_*i*_{*S*_*i*_ −*S*_*i*+1_}, in which for each model *S*_*i*_ −*S*_*i*+1_ is the number of new infecteds on day *i*. The basic reproduction number, written ℛ_0_, is the special case for infection seeded into a fully susceptible population (*S*_*i*_ ≈ *N*) and prior to any mitigation (*α*_*i*_ = 1).

By design, the *SI*_*t*_*D* model explicitly incorporates the infectiousness profile, {*β*_*j*_}, and the infection-to-death distribution 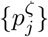, such that these can be chosen according to clinical data. For the other models in which these aren’t explicit, it is helpful to understand what are the implied {*β*_*j*_} and 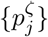. Calculation of these is in A.

#### 2.2.6 Model initial conditions

In this paper, *i* indexes the number of days since 15 Feb 2020, which is day *i* = 0. We assume initially zero deaths, *D*_0_ = 0, and zero recovereds, *R*_0_ = 0 (for models including *R*) and *S*_0_ = *N* − *I*_0_ for parameter *I*_0_ which is to be inferred. For the infected-age-structured model, it is necessary to specify how the *I*_0_ initial infecteds are distributed by infected age {*I*_0,*j*_}. When the epidemic is growing exponentially the distribution of infecteds by infected age converges to an equilibrium distribution (see B), thus we assume assume convergence to this equilibrium distribution in the dynamics prior to day *i* = 0, and for the numerical calculations in this paper we use the equilibrium distribution as the initial condition at *i* = 0.

### 2.3 Observation model

The epidemic models above are deterministic. It is common to account for variability in the observed number of daily deaths, 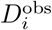, on day *i*, in mechanistic models of infectious disease transmission via a negative binomial observation model (e.g. Mathews et al., 2007; Cauchemez and Ferguson, 2008). The negative binomial model admits overdispersion, which is often present in count data on cases or deaths from an infectious disease owing to spatial and demographic heterogeneities, or other unmodelled processes (Held et al., 2019). We adopt the negative binomial model, assuming that

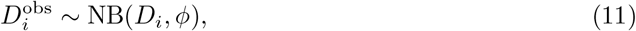

independently for each *i*, where NB(*μ, ϕ*) denotes the negative binomial distribution with mean *μ* and dispersion parameter *ϕ* defined such that the variance equals *μ* + *ϕμ*^2^ (Robinson and Smyth, 2008).

### 2.4 Inference methods

We denote by Θ^∗^ the free parameters that appear in the respective dynamical models, such that *D*_*i*_ = *D*_*i*_(Θ^∗^), and by Θ = (Θ^∗^, *ϕ*) the vector of all the parameters including in the observation model (11). The data 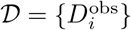 are the daily deaths indexed by time *i*. We denote by *P*(𝒟|Θ) the likelihood function for Θ under observation model (11). When adopting a Bayesian approach and specifying a prior distribution *P* (Θ) on Θ, the posterior distribution for Θ is then

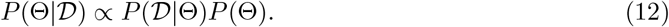

In the results sections below, where we compute point estimates of Θ we do so using the *maximum a posteriori* (MAP) estimator

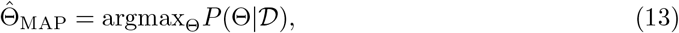

then the inferred dynamics, and the epidemiological parameters inferred from them (defined in §2.2.5), are based on solutions of the respective epidemic model with 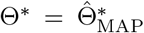. The MAP estimator corresponds to the widely used maximum-likelihood estimator (MLE) 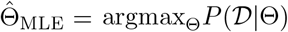 if the priors are uninformative, or more generally if *P* (Θ) is constant on a domain that contains 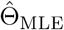 and zero elsewhere. Elsewhere we target the full posterior distribution (12) by sampling from it using Markov chain Monte Carlo (MCMC). We used an adaptive Metropolis-Hastings MCMC algorithm (Haario et al., 2001) which involves, after a warm-up phase, adapting the covariance matrix of a multi-variate Gaussian proposal distribution according to the covariance of the accepted samples. Convergence to the stationary distribution is hastened with an initial maximisation of the posterior density with respect to Θ using a covariance matrix adaptation evolution strategy (CMA-ES) (Hansen et al., 2003). These methods were implemented using the PINTS python package (Clerx et al., 2019).

For priors on Θ, the priors arising from the clinical data are detailed in Table 1 and §2.1.3, in addition to which we specify: *ρ* ∼ *U* (1, 10) and *I*_0_ ∼ *U* (1, 5 × 10^7^) consistent with wide range of possible values for ℛ_0_ and number of initial infecteds; *t*^∗^ ∼ 𝒩 (31, *σ* = 3), where *i* = 31 corresponds to where the Google mobility data, shown in Figure 1A, first suggest a substantial reduction in mobility; and *α*_*b*_ ∼ *U* (0, 1) and *ϕ* ∼ *U* (0, 1) such that both are uniform on their possible ranges of values. For the *SIR*-type models, transition rates in equations (4), (7) and (8) have upper limits such that, for example, no more than the entirety of a compartment can transition out in a single time step. Consequently, we take *θ* ∼ *U* (0, 1) for the *SIRD, SIRD*Δ*D & SIURD* models, *ξ* ∼ *U* (0, 1) for the *SIURD* model and *θ, η, ξ* ∼ *U* (0, 0.5) for the *SE*^2^*I*^2^*U* ^2^*RD* model. Due to the additional scaling by 1*/θ* in the relationship between R_*i*_ and *ρ* for SIR-type models (equation 10) plausible values for *ρ* are contracted and we use *ρ* ∼ *U* (0, 3).

## 3. Results

### 3.1 A change in mixing intensity, and not ‘herd immunity’, is necessary to explain the epidemic dynamics

A theory after decline from the initial epidemic peak was that the epidemic dynamics were affected little by NPIs and could be be explained by ‘herd immunity’ (Lourençco et al., 2020), that is, that R_*i*_ in (9) had reduced to below 1 because the number of remaining susceptibles, *S*_*i*_, had sufficiently decreased to curtail the growth. The theory that depletion of susceptibles is adequate to explain the dynamics can be tested by fitting both models to the data; the ‘herd immunity’ theory corresponds to the a special case of the general model that has a step change in *α*_*i*_ but with the restriction of having constant *α*_*i*_ = 1, achieved in the model by setting *α*_*b*_ = 1. This supposes no effect from NPIs, and thus that the decline in the epidemic must be on account of the depletion of susceptibles. In this restricted model the parameter *t*^∗^ is redundant, therefore the restricted model has two fewer parameters than the unrestricted model. To consider this, we use the *SI*_*t*_*D* model, with the infectiousness profile and infection-to-death distribution parameters fixed to the values shown in Table 1, and noninformative priors on *ρ, I*_0_, *t*^∗^, *α*_*b*_, *δ*, and *ϕ*, then fit the model to the the England and Wales deaths data described in §2.1.1. We then do likewise for the restricted model, with the extra restriction that *α*_*b*_ = 1 and with *t*^∗^ removed.

The fitted models are shown in Figure 3, from which it is clear that the full model matches well to the data but the restricted model matches very poorly. The difference in the value of the maximised values of the log-posterior, log *P* (Θ | 𝒟), for the two models is 260, which for models differing, as here, by two degrees of freedom is overwhelming evidence that the restricted model is inadequate.

Some values of epidemiological parameters in the fitted restricted model also seem implausiable; for example, the fitted *I*_0_ is greater than 1 million people, and the IFR is ∼ 4 times smaller than the best estimate from O’Driscoll et al. (2020) — see Figure 3 for all of the inferred parameter values.

The values of the maximised likelihoods, and visual inspection of the fits, make clear that a change in transmission dynamics over time, via *α*_*i*_ in the model, is necessary to explain the epidemic dynamics during the first outbreak. In other words, it was never plausible that ‘herd immunity’ was responsible for the end of the first outbreak, as confirmed by the subsequent resurgence of infections in autumn of 2020.

### 3.2 Inference for epidemiological parameters is unreliable if based on non infected-age structured models

To investigate the impact of model error on the values and uncertainties of inferred parameters — and in particular whether simpler epidemic models such as *SIRD, SIRD*_ΔD_, *SIURD* and *SE*^2^*I*^2^*U* ^2^*RD* can be used to infer accurately the epidemiological parameters — we generate synthetic data from the time-structured SI_t_D model (§2.1.1) and observation model (11), with the model parameters as specified in Table 2, then refit both the SI_t_D and those other models to compare the inferred parameter values with the true ones.

**Table 2:**
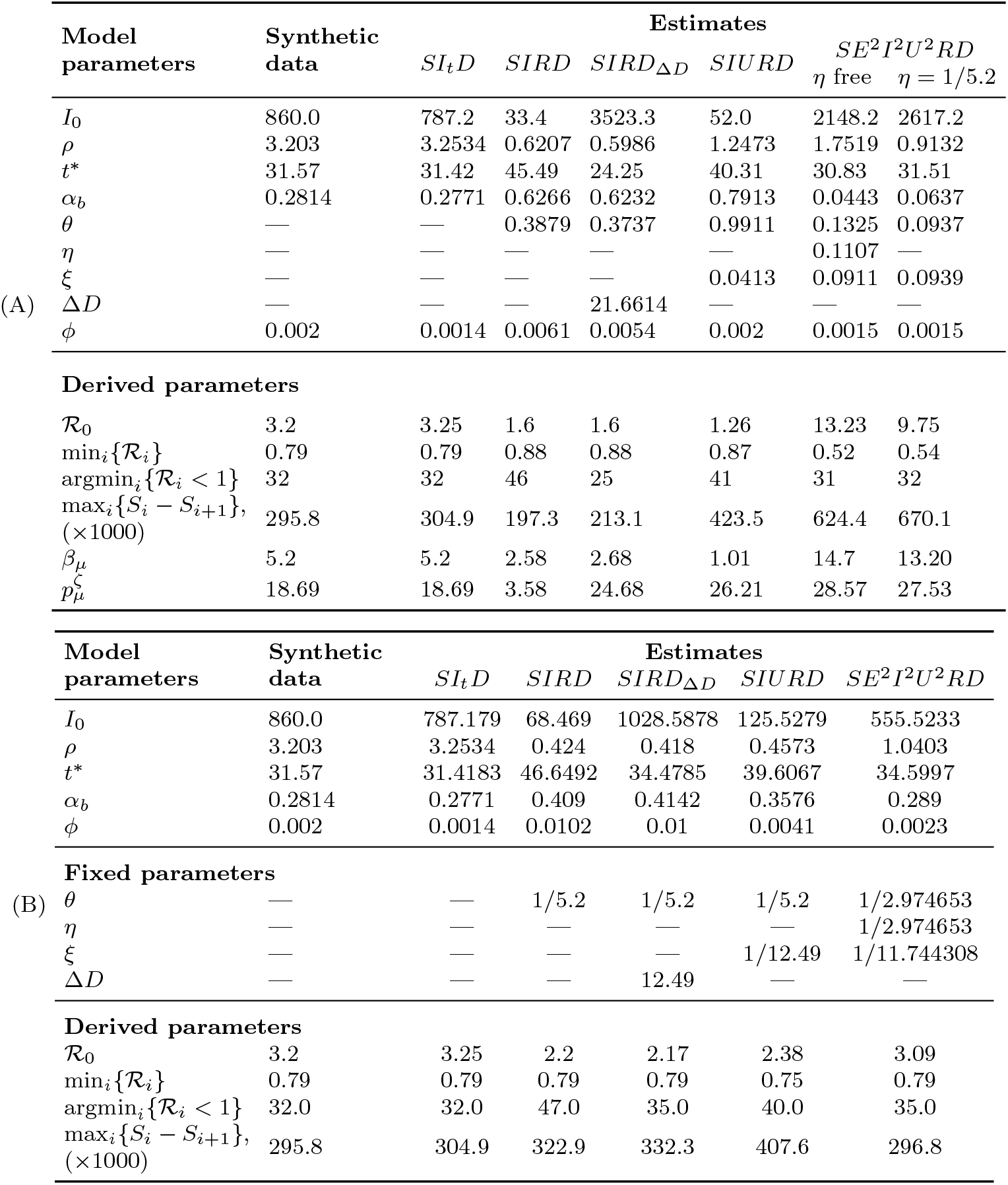
Details of the simulation study described in section §3.2, including parameter values, in addition to those in Table 1, used to create synthetic data from the *SI*_*t*_*D* model; and MAP estimates of the model parameters and derived epidemiological parameters from fitting the various models to the synthetic data. The (–) indicate parameters not relevant to the particular model. The derived parameters are: the basic reproduction number R_0_; the minimum value of ℛ, min_*i*_{ℛ_*i*_}; the index of the day at which ℛ_*i*_ first decreases below 1, argmin_*i*_{ℛ_*i*_ *<* 1}; the maximum number of new infections on any day, max_*i*_ {*S*_*i*_ −*S*_*i*+1_} ; the mean, *β*_*μ*_, of the infectiousness profile; and the mean of the infection-to-death distribution, 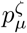. The (A) and (B) tables differ in that (B) involves extra parameters being fixed when the models are fitted, as described in §3.2.

We fit the models, fixing the IFR, *δ*, to the default value from Table 1 and treat the other parameters as free. Hence the free parameters include those that determine the implied infectiousness profile and the infection-to-death distributions. The data-generating and fitted parameters values are summarised in Table 2A, and results are plotted in Figure 4.

**Figure 4:**
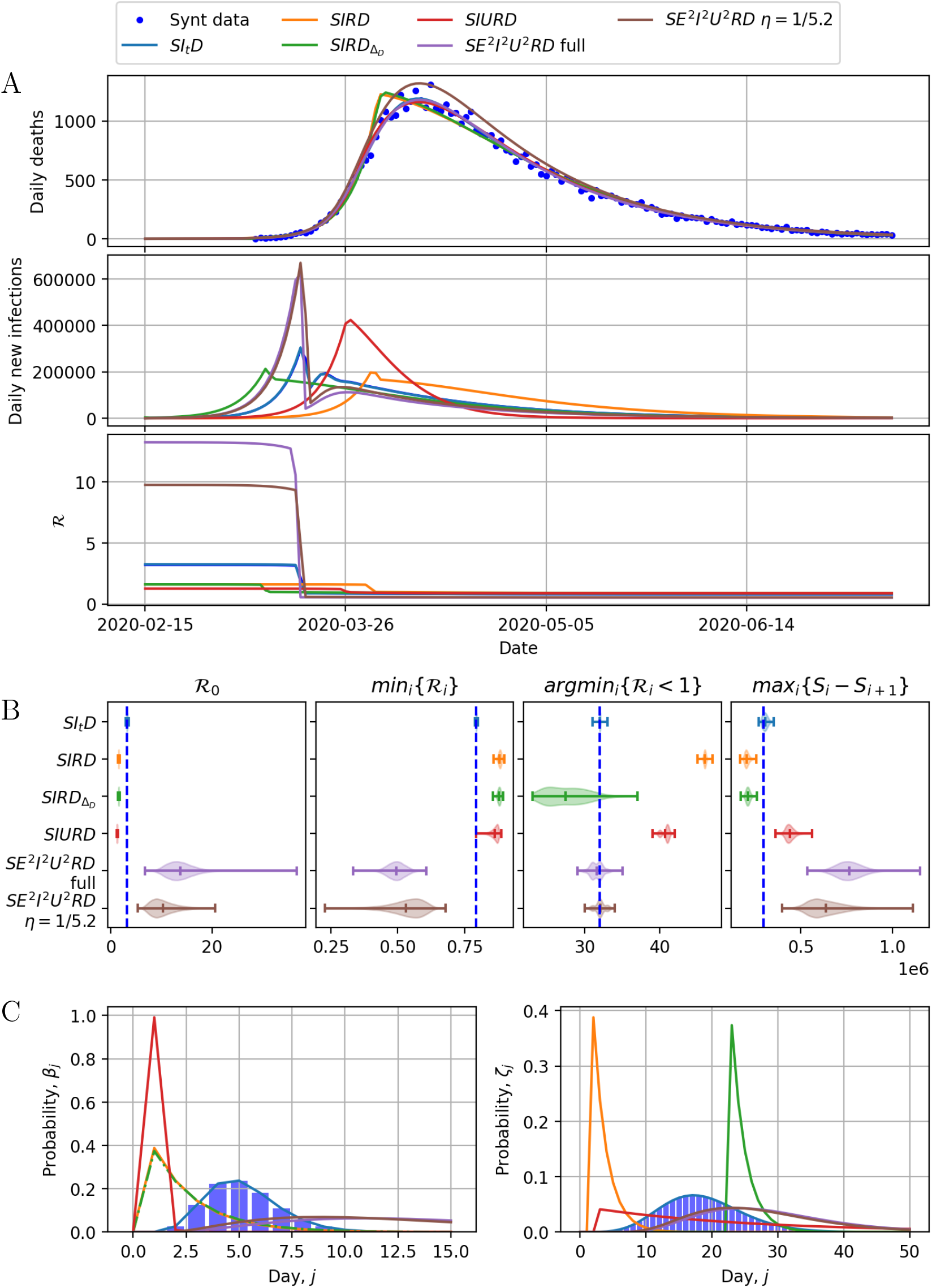
(A) Synthetic data from the *SI*_*t*_*D* model generated using parameters in Tables 1A and 2, and dynamics of fitted *SI*_*t*_*D, SIRD, SIRD*_Δ*D*_, *SIURD* and *SE*^2^*I*^2^*U* ^2^*RD* models, corresponding to MAP estimates of parameters per Table 2A. (B) Violin plots of the posterior marginals for derived epidemiological parameters under the different models, with dashes representing (min, mean, max) and the vertical blue line showing the true values. (C) Infectiousness profiles and infection-to-death distributions for the fitted models.

Figure 4A shows that each fitted model matches broadly well the deaths data. The *SIRD* and *SIRD*_ΔD_ models provide similar and relatively reasonable fits to the synthetic death data, but the peak daily deaths in these model fits occurs almost one week before the true peak.

Each of the simpler models — *SIRD, SIRD*_Δ*D*_, *SIURD* — substantially underestimate ℛ_0_ compared to the true value, in each case at least by a factor of 1.6, but estimates from *SEI*^2^*I*^2^*U* ^2^*RD* are vastly too large and with very high posterior variance; see Fig 4B. The latter also had the largest number of free parameters, so we also considered the case with *η* fixed to the value 1*/*5.2, which is an appropriate choice for this parameter in the sense that it fixes the model’s onset-to-infection interval to match with the mean we assumed when setting the default infection-to-death distribution (see Section 2.1.3). Even with this parameter value fixed, the there is little improvement in the inferred values for ℛ_0_. During the epidemic decline after introduction of NPIs, the misspecification of the model is less impactful on the inferred values for ℛ_*i*_, but the inferred time, *t*^∗^, at which NPIs are inferred to impact mixing intensity is highly variable between models: for example it is inferred around 14 days too late for the *SIRD* model, 7 days early for *SIRD*_ΔD_ and 9 days late for *SIURD* (Table 2). Compared to the data-generating model, the inferred peak of new infections, max_*i*_*I*_*i*,1_, and the inferred timing of this peak, argmax*iI*_*i*,1_, are also very inaccurate. For *SIRD* and *SIRD*_ΔD_ models the peak is approximately 36% too low and the timing is respectively 14 days late and 2 days early. For the SIURD model the inferred peak is approximately 43% too high and 9 days too late. Because here we are fixing the IFR and are fitting to death data, all model versions return approximately the same remnant susceptible pool (*S*_*i*=end_*/N* ≈ 0.88) by design. This means that from equation (9) if mixing intensity returns to pre-epidemic levels following the first wave (*α*_*i*_ = 1), and all other parameters are fixed, then ℛ_*i*_ ≈ 0.88 ℛ_0_. This provides an upper bound on the median _*i*_ for a subsequent unmitigated outbreak equal to 2.8 in this simulated example. Because the simple SIR-like models underestimate ℛ_0_, they also substantially underestimate ℛ_*i*_ for a subsequent unmitigated outbreak, suggesting an upper bound of the median ℛ_*i*_ ≈ 1.4 (*SIRD, SIRD*_Δ*D*_) or 1.1 (*SIURD*) which are highly misleading. In contrast, estimates of the median unmitigated ℛ_*i*_ for the *SEI*^2^*I*^2^*U* ^2^*RD* models are vastly inflated. In the context of influencing policy-making, the errors arising from the simple SIR-like models are unacceptably large. Figure 4c shows that the implied infectiousness profile and infection-to-death distributions for the fitted models were all quite different to the true ones.

In the preceding investigation with synthetic data, the data-generating *SI*_*t*_*D* model was at an advantage over the other models through having its infectiousness profile and infection-to-death distribution specified correctly, and without free parameters related to these that needed estimating. To what extent did the other models perform poorly on account of their extra parameters, rather than their model misspecification? To investigate, we fixed the values of those parameters to “optimal” values in the sense of matching moments of their implied infectiousness profile and infection-to-death distribution matched as closely as possible to the true ones, and with the consequence that the number of free parameters is then the same for each model, including for the data-generating *SI*_*t*_*D* model. The parameters are summarised in Table 2B and explained as follows.

To match the mean of the infectiousness profile to the true one (recalling that the relationship between this profile and the model parameters for the various models is explained in A), for *SIRD, SIRD*_Δ*D*_ and *SIURD* models we take the infectivity rate *θ* = 1*/*5.2 where, per Table 1, 5.2 days is the mean of the assumed infectiousness profile. For the infection-to-death distribution, matching to the mean of 18.69 days for *SIRD*_Δ*D*_ entails taking entails taking Δ_*D*_ = 18.69 −1 − 5.2 = 12.49 and for *SIURD* taking *ξ* = 1*/*12.49 respectively. For the *SE*^2^*I*^2^*U* ^2^*RD* model there are many choices for *η* and *θ* that yield an implied infectiousness profile (A) with appropriate mean, however in general the variance is higher than that for the clinical infectiousness profile (§2.1.3). We therefore fix *η* = *θ*, which minimises the variance in the implied infectiousness profile. We then select fixed values for *θ & ξ* to match the means of the clinical infectiousness and time-to-death profiles to those of the implied profiles.

The results of this second fitting, with extra fixed parameters, are shown in and the results are shown in Table 2B and Figure 5. For *SE*^2^*I*^2^*U* ^2^*RD* in particular the results are much improved. Figure 5B shows that the derived epidemiological parameters are much more reliably inferred, although ℛ_0_ is still slightly underestimated and argmin_*i*_ {ℛ_*i*_ *<* 1} overestimated. The better performance is because the the inclusion of the E state in this model, with the duplicated compartments for E, I and U, enables the fitted infectiousness profile and infection-to-death distribution to match reasonably closely to the true ones, as shown in Figure 5c. For each of the other models this is not the case, there is a much larger mismatch, as shown in Figure 5c, and a consequence is that the values of inferred epidemiological parameters, particularly ℛ_0_, remain highly unreliable.

**Figure 5:**
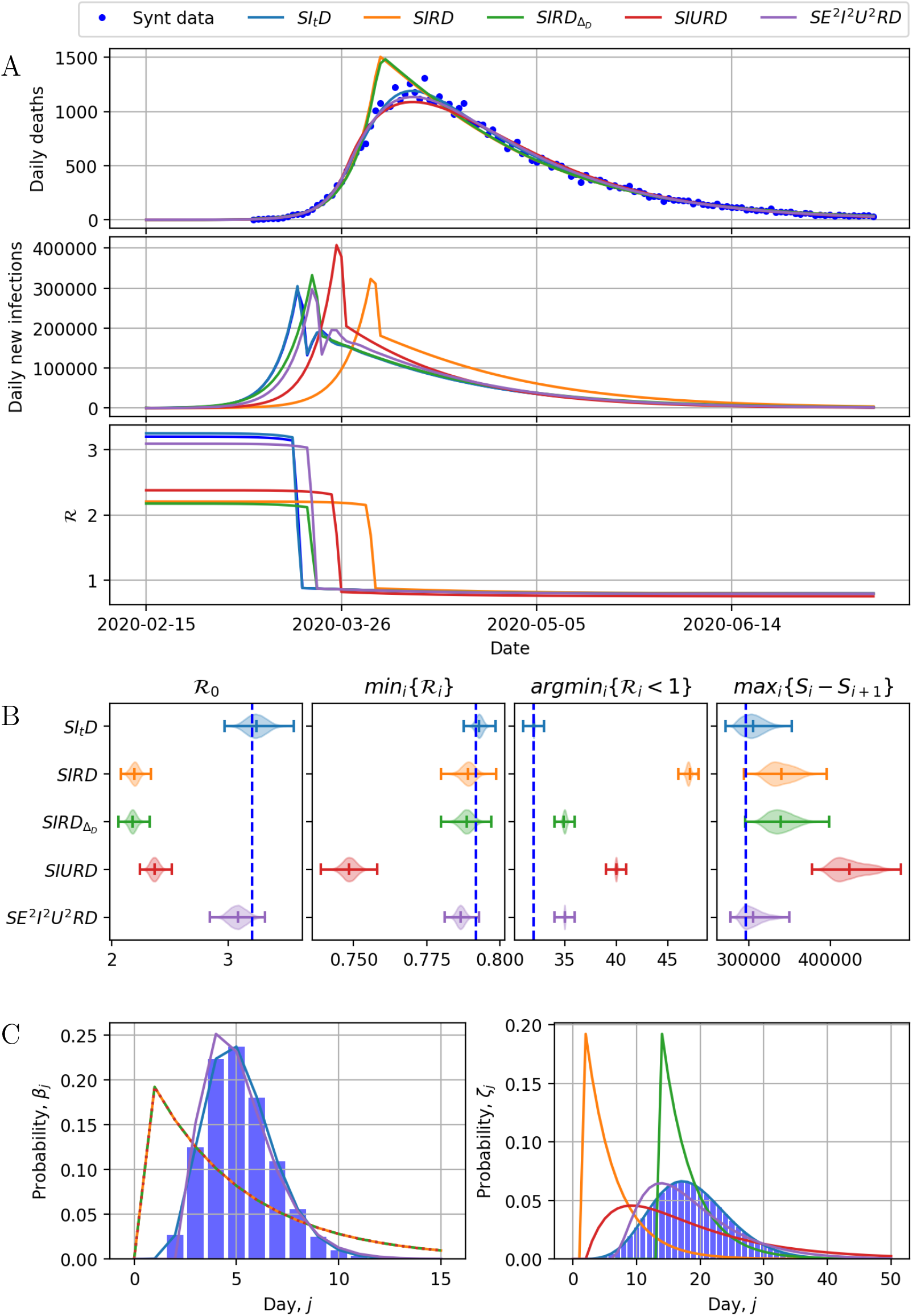
(A) Synthetic data from the *SI*_*t*_*D* model (§2.2.1) generated using parameters in Tables 1 and 2; and dynamics of fitted *SI*_*t*_*D, SIRD, SIRD*_Δ*D*_, *SIURD* and *SE*^2^*I*^2^*U* ^2^*RD* models for the parameters indicated in Table 2B; this version has additional fixed parameters, whose values are chosen *a priori* such that for each model the infectiousness profile and infection-to-death distribution, shown in (C), matches as closely as possible to the data-generating ones. (B) Violin plots of the derived epidemiological parameters uncertainties for each model and the synthetic data true values (blue dotted lines).

In summary, a model such as *SE*^2^*I*^2^*U* ^2^*RD* could plausibly lead to reasonable inference for epidemiological parameters, provided the number of pre-infectious, infectious and post-infectious compartments is judiciously chosen such that the implied infectiousness profile and infection-to-death distribution can match suitably closely the clinical data; though if, as in this paper, inference is based on deaths data (§2.1.3) then it appears important that the parameters of such a model are estimated *a priori* from the clinical data.

The results in Table 2 & Figures 4 and 5 are generated from a single synthetic data set; however, the results are representative of the large number repetitions performed.

### 3.3 Bayesian inference leads to small posterior uncertainty when conditioning on the infectiousness profile and infection-to-death distribution

In this section, we adopt a Bayesian approach to characterise the uncertainty in the inferred parameters and predictions. We fixed the infectiousness profile and infection-to-death distribution to their default values from Table 1. The priors for the other parameters are as specified in §2.4, and results for fitting the *SI*_*t*_*D* model are shown in Figure 6. Figure 6A shows bi- and uni-variate marginal posterior distributions for the model parameters; these suggest relatively small posterior variance, and small posterior covariance except between *ρ* and *α*_*b*_; and *I*_0_ and *α*_*b*_. Figure 6B shows a good match between the deaths data and the model dynamics based on the maximum a-posteriori (MAP) model parameter values. Figure 6C shows corresponding inferred new infections, which peak in mid-March, owing to the inferred sharp change in *α*_*i*_, then decline. Posterior distributions for derived epidemiological parameters are shown in Fig 6E. The posterior for ℛ_0_ is concentrated in the region ∼3 to 3.5; min_*i*_{R_*i*_} is concentrated around 0.795; almost all the posterior mass for argmin_*i*_{ℛ_*i*_ *<* 1} is on *i* = 32, which is 18th March; and for max_*i*_{*I*_*i*,1_} there is somewhat more posterior uncertainty, with values between ∼220,000–400,000 plausible, and the posterior mode at ∼280,000.

**Figure 6:**
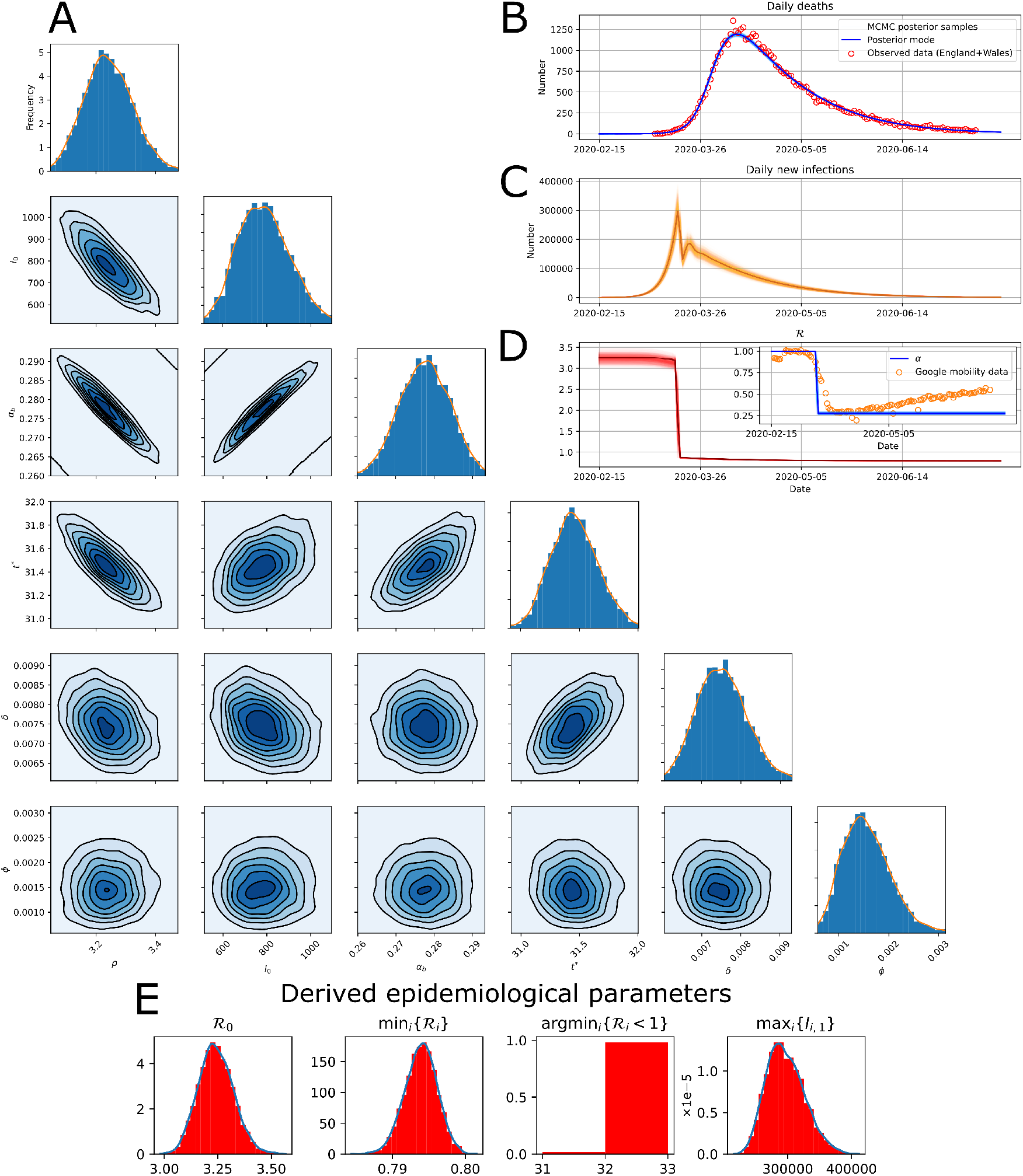
Results from the Bayesian inference described in §3.3. (A) Uni- and bivariate marginal posterior distributions for the model parameters. (B) Daily death data, with superimposed lines showing modelled deaths from 1000 posterior samples (transparent) and MAP parameter values (solid). (C) Inferred daily new infections based on MAP parameter values. (D) Posterior MAP estimate of ℛ_*i*_, with inset showing correspondence between inferred *α* and average of work-place and transit Google mobility data. (E) Posterior distributions for derived epidemiological parameters.

The change in *α*_*i*_ matches quite closely to the Google mobility data, both in the timing, *t*^∗^, and the extent of the drop, *α*_*b*_, as shown in Figure 6D. These numbers are consistent with a reduction in mixing throughout March (before the official lockdown was mandated) due to voluntary changes in mixing intensity, perhaps driven by media coverage and government announcements prior to lockdown. In other words, the results suggest the Google mobility data may accurately reflect the timing and drop in population mixing around the period a lockdown was mandated.

Our inferred values of ℛ_0_ are slightly higher than estimates based on an exponential growth model fitted to the death data [2.6 (2.4–2.9), Lonergan and Chalmers (2020)], but consistent with other estimates for the UK (Royal Society SET-C, 2020). Estimates of ℛ_*i*_ after lockdown are consistent with those from exponential decay models fitted to death data (Lonergan and Chalmers, 2020), and real-time estimates based on reported cases in the period April-June 2020 [see Royal Society SET-C (2020) and references therein] or surveyed contact rates in late March 2020 (Jarvis et al., 2020).

### Incorporating uncertainty in the infectiousness profile and infection-to-death distribution leads to misleading conclusions owing to model error

Here we repeat the analysis of the previous section, except this time incorporating uncertainty on the four parameters—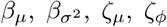—describing the infectiousness and infection-to-death distributions. The priors on these parameters are as described in §2.1.3 and summarised in Table 1. Results analogous to Figure 6 are shown in Figure 7. These results show that posterior variances are somewhat larger but not substantially so, although MAP estimates are considerably different.

**Figure 7:**
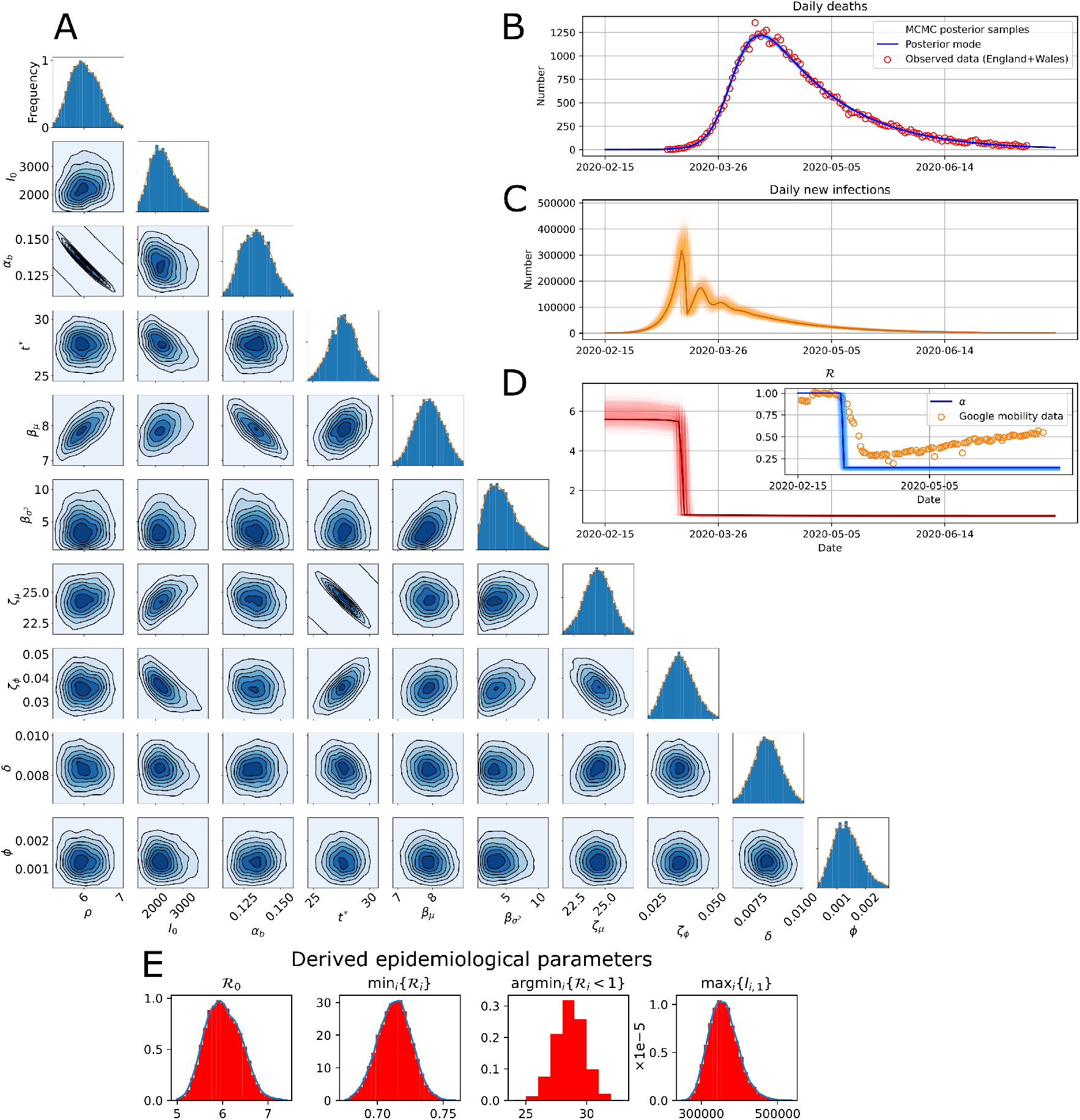
Results from the Bayesian inference described in §3.4. The interpretation of (A– E) are as for Fig 6. These results are analogous to those in Fig 6, except here the analysis involved placing priors on—rather than conditioning on—the parameters 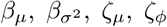 of the infectiousness profile and infection-to-death distribution.

The MAP parameter gives visually a similarly good fit between model and observed data (Figure 7B), predicting daily new infections to peak in early- to mid-March, after which a sharp drop in new infections is observed (Figure 7C). The inferred mixing intensity in this case still agrees reasonably with the UK Google mobility data (Figure 7D), but is predicted to drop earlier and further than was inferred in §3.3, when the values of those four parameters were instead conditioned upon.

The posteriors for derived parameters show ℛ_0_ is distributed largely between 5 and 7 and peaking at ∼ 6; min_*i*_{ℛ_*i*_} is concentrated around 0.72, argmin*i*{ℛ_*i*_ *<* 1} is some time between 11th and 17th March, with posterior peaking on 14th March, and max_*i*_ {*I*_*i*,1_} somewhere between ∼ 250,000–500,000, peaking at ∼ 350,000 (Figure 7E). These posterior distributions are somewhat in conflict with some informative priors (the influence of the priors can be seen in Supplementary Figure S1). For example, the peak infectiousness occurs ∼ 3 days later than estimated by Ganyani et al. (2020), and an _0_ of around 6 is higher than most very early estimates of ℛ_0_ (Park et al., 2020). However, reproduction numbers of around 6 have been reported for other European countries (Billah et al., 2020).

A possible explanation for the discrepancy in results between this section and §3.3, and with estimates from clinical data such as Ganyani et al. (2020), is that it is a consequence of model error. To investigate this, we repeated the Bayesian inference of the present section and of §3.3 for the synthetic data shown in Figure 4. Doing so leads to posterior distributions that are consistent with the data-generating parameters, and consistent with each other, with larger variance under the approach of this section in which there is prior uncertainty on the four additional parameters. Results for these cases are in the Supplementary Material (Supplementary Figures S2 and S3). In other words, the results of the Bayesian inference are as we might expect when the data arise from the assumed model, supporting the possibility that the unexpected results for the real data are on account of model error.

If so, what are the possible sources of model error? The negative binomial observation model heavily penalises discrepancies between modelled and observed deaths when the number of daily deaths is small, especially when the over-dispersion parameter, *ϕ* is small. The MAP estimate of *ϕ* is indeed small—around 10^−3^—especially so with the extra four degrees of freedom considered in this section, which enable the deterministic part of the model to explain more of the variability in the data. A consequence is that the inference is dominated by the model having to match well the data in the period when the number of daily deaths is small but growing rapidly. This is the period when the deterministic component of the model, which assumes a homogeneously mixing population and neglects stochastic effects in the epidemic process, describes the dynamics least well. In summary: enabling the extra four degrees of freedom in this section appears to lead to an overfitted model, which leads to underestimation of the overdispersion parameter (and variance). The posterior consequently has low variance, but with inference dominated by the dynamics in a period when the model is likely to be least accurate, and the low-variance posterior is located in a region of parameter space that suggests model error is making the results unreliable.

The Supplementary Material contains results of some further exploration of this issue, including where we have fixed *ϕ* at two larger values, 0.01 and 0.1, (Supplementary Figures S4 and S5), which inflates posterior variance and shifts the MAP estimates to more plausible values, e.g. 4.5 for ℛ_0_; and a case with the Negative Binomial observation model replaced by a Gaussian model (Supplementary Figure S6), such that the inference is not dominated by the early period with low prevalence, and for this case again the MAP estimates of parameters are more in accordance with other sources and with the results of §3.3.

## 4 Summary and discussion

We have explored various simple epidemic models, particularly investigating the importance of accurately characterising the infectiousness profile and the infection-to-death time distribution. These distributions are well characterised in the clinical literature and easy to incorporate into an SIR-type model structured by infected age. The results show that it is essential to incorporate them in models used for inferring the underlying epidemic dynamics from death data. Basic SIR models implicitly mis-specify the infectiousness and infection-to-death distributions and lead to highly misleading inferences about the impact of NPIs, the time and magnitude of peak infections, and the basic reproduction number.

In the particular context of the spring 2020 SARS-CoV-2 outbreak in England and Wales, we used the infected-age-structured model, *SI*_*t*_*D*, to compare the hypothesis that the epidemic decline was on account of NPIs versus the hypothesis, which was entertained at the time, that the decline was owing to ‘herd immunity’. When using distributions for the infectious profile and infection-to-death distribution that are drawn from clinical literature, but fitting all the other model parameters, we found very strong evidence in favour of efficacious NPIs, rather than herd immunity, as the explanation for the epidemic decline (Figure 3). This conclusion was in spite of accommodating the possibility of an implausibly low IFR, *δ*, and high initial number infected, *I*_0_, both of which are to the benefit of the herd immunity hypothesis. This indicates that the death data, combined with infectiousness profile and infection-to-death distributions available from clinical data, enable dismissing of the ‘herd immunity’ hypothesis, without needing to know the IFR very accurately.

The limitations of very basic SIR-type models in mis-specifying the infectiousness profile and the infection-to-death has been pointed out by Keeling and Rohani (2008); Wearing et al. (2005). Our work extends this to consider the impact of mis-specification when the model is used to infer epidemiological parameters and dynamics from data, and particularly when there is an abrupt change in population mixing intensity. Our findings show major errors in the inferred dynamics—for example, timing of changes in population mixing can be wrong by weeks— using basic SIR-type models, suggesting that untangling the impact of NPIs adopted in quick succession (e.g. Dehning et al., 2020) may be especially error-prone if using a simple SIR-like model. Models such as *SE*^2^*I*^2^*U* ^2^*RD* that incorporate multiple infected states (in this case preinfectious, infectious, post-infectious) and multiple sub-compartments per state can potentially perform well if the number of states, the number of sub-compartments, and the values of rate parameters are suitably chosen, but in our view this entails more complexity and no advantage compared using the *SI*_*t*_*D* model.

Our estimates for ℛ_0_ are significantly larger when we take a fully Bayesian approach and fit parameters describing the infectiousness profile and infection-to-death distribution. Much of the variation between early estimates from case data has been attributed to assumptions for the generation time distribution (Park et al., 2020), which is closely linked to the infectiousness profile. The need to quantify uncertainty in estimates of ℛ_0_ for use in policy-making has been highlighted, for example by Royal Society SET-C (2020). Uncertainty analysis of a spatial agent-based model suggests that parameters that largely determine the infectiousness profile (exposure to onset delays, onset to isolation delays) remain significant sources of overall uncertainty (Edeling et al., 2021). We found that incorporating uncertainty by placing priors with relatively large variance on the four parameters characterising the infectiousness profile and infection-to-death distribution — as opposed to conditioning on them — yielded low-variance posteriors concentrated in implausible regions of parameter space, contrary to our initial expectation that the additional prior uncertainty would largely just inflate the posterior variance. We understand this to be the impact of model error, with the negative binomial observation model leading to inference being dominated by a period in which the deterministic dynamics are least reliable. A tendency of deterministic models to yield overconfident and error-prone estimates of ℛ_*i*_ from early incidence data has also been documented (King et al., 2015).

We have argued in favour of an SI_*t*_D model structured by infected age over a basic SIR model, especially when the goal is inference from death data, but this model is itself only a coarse population-level model with many limitations. The model assumes homogeneous mixing, and neglects stochastic effects which might be appreciable when prevalence is low. To keep the model simple, we accommodated only a step change in the population mixing intensity, *α*_*i*_. For the observation model for deaths, we assumed a negative binomial model and conditional independence between different days, which are strong parametric assumptions upon which the inference can be sensitive. We did not attempt to model subtleties such as how hospital fatality rates may depend on case loads within hospitals (Docherty et al., 2020). Hence there are many possible sources of model error. In the context of inference with this model, we have found that affording too much freedom through high prior variance on parameters enables overfitting of the deterministic part and amplifies the impact of model error. Erring on the side of choosing priors with high variance is therefore not necessarily a conservative strategy, at least when model error is non-negligible.

## Data Availability

Open source software to reproduce the results in this paper is available at
https://github.com/DGWhittaker/nottingham_covid_modelling

## Software Availability

Open source software to reproduce the results in this paper is available at https://github.com/DGWhittaker/nottingham_covid_modelling.

## Funding

This work was supported by the Wellcome Trust [grant number 212203/Z/18/Z]. DGW, ADH-R and GRM acknowledge support from the Wellcome Trust via a Senior Research Fellowship to GRM. KJB acknowledges support from a University of Nottingham Anne McLaren Fellowship. This research was funded in whole, or in part, by the Wellcome Trust [212203/Z/18/Z]. For the purpose of open access, the author has applied a CC-BY public copyright licence to any Author Accepted Manuscript version arising from this submission.

## Acknowledgements

We thank Frank Ball and Theodore Kypraios for helpful discussions.

## A Infectiousness profiles and time-to-death distributions for *SIRD, SIRD*_Δ*D*_ and *SE*^2^*I*^2^*U* ^2^*RD* models

For our models without a latent state defined by equations (4) & (7), the proportion of newly infected hosts remaining in an infectious state (*I*) at infected-age *j* is simply those who are not removed by the previous time-step *A*_*j*_ = (1 −*θ*)^*j*−1^, and *β*_*j*_ = *θ*(1 −*θ*)^*j*−1^. The probability mass function for death (as before conditioning on death as the outcome of the infection) can be calculated by summing over the probabilities of Markov chains that arrive in *D* at infected-age *j*. For the *SIRD* model:

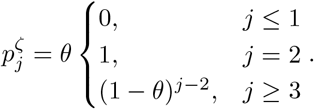

The distribution of arrival times in *D* is shifted for the *SIRD*_Δ*D*_ model,

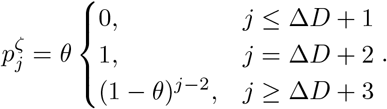

For the *SIURD* model:

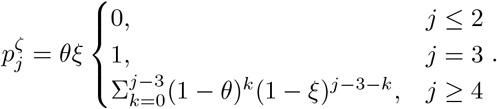

For the slightly more complicated *SE*^2^1^2^*U* ^2^*RD* model described in equation (8) we construct a matrix describing the Markov transitions following infection (Diekmann et al., 2021)

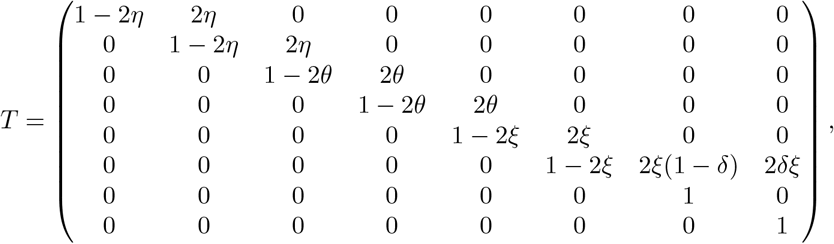

where *T*_*n,k*_ is the probability of transitioning from state *n* to state *k*. Describing a newly infected individual in *E*_1_ by a state vector *i*_0_ = (1, 0, 0, 0, 0, 0, 0, 0), the proportion of new infections in *I*_1_ at infected-age *j* is 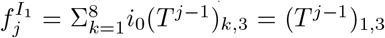, where *T*^*j*^ indicates the *j*th power of *T*. Similarly 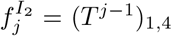. Hence *A*_*j*_ = (*T*^*j*−1^)_1,3_+ (*T*^*j*−1^)_1,4_. If 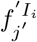 denotes the proportion of infections in *I*_i_ at time-step *j*^′^ after arriving in *I*_*i*_, then 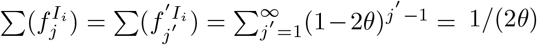, and thus *β*_*j*_ = *θ*((*T*^*j*−1^)_1,3_ + (*T*^*j*−1^)_1,4_). The probability of death at infected-age *j* is 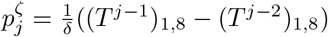.

## B Pseudo-steady distribution of infecteds by infected age

During a phase of epidemic growth with *S*_*i*_ ≈ *N* and *λ*_*ij*_ = *λ*_*j*_ constant with respect to *i*, the distribution of *I*s by infected age reaches a dynamic equilibrium,

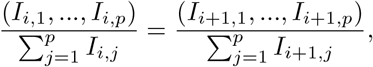

and the number of infecteds grows exponentially,

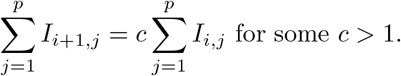

Together these imply *I*_*i*+1,*j*_ = *cI*_*i,j*_. Then under model (1), we get,

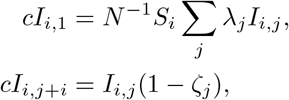

and the solution of these equations for {*I*_*i,j*_}, after eliminating *c*, is the equilibrium distribution.

## Supplementary Figures

**Figure S1:**
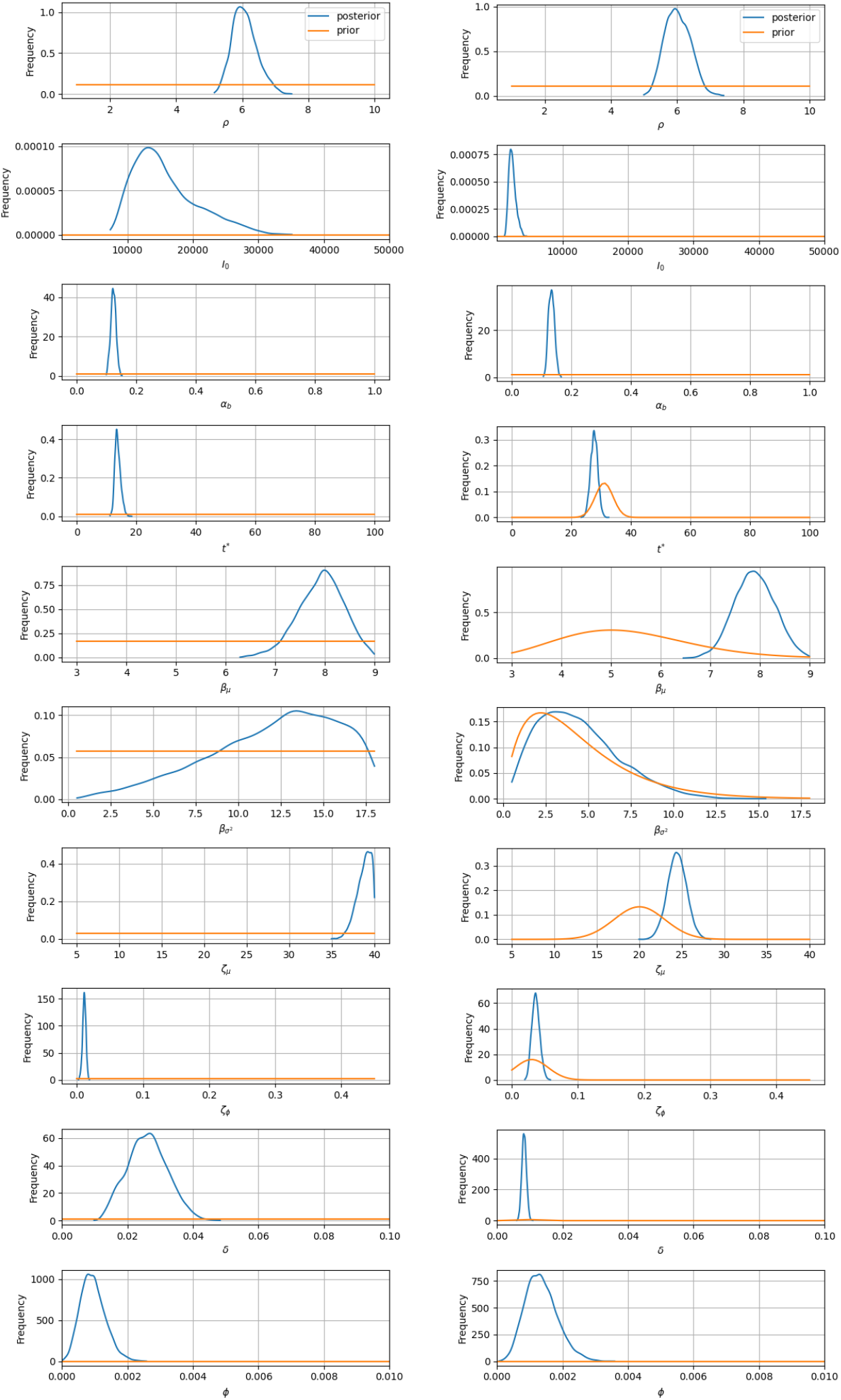
Comparison of posterior distributions from the ‘fully’ Bayesian inference in which the priors are diffuse (left) and informative (right). In each case, the posteriors (blue) are overlaid on the priors (orange). The right panel corresponds to results shown in Figure 7.

**Figure S2:**
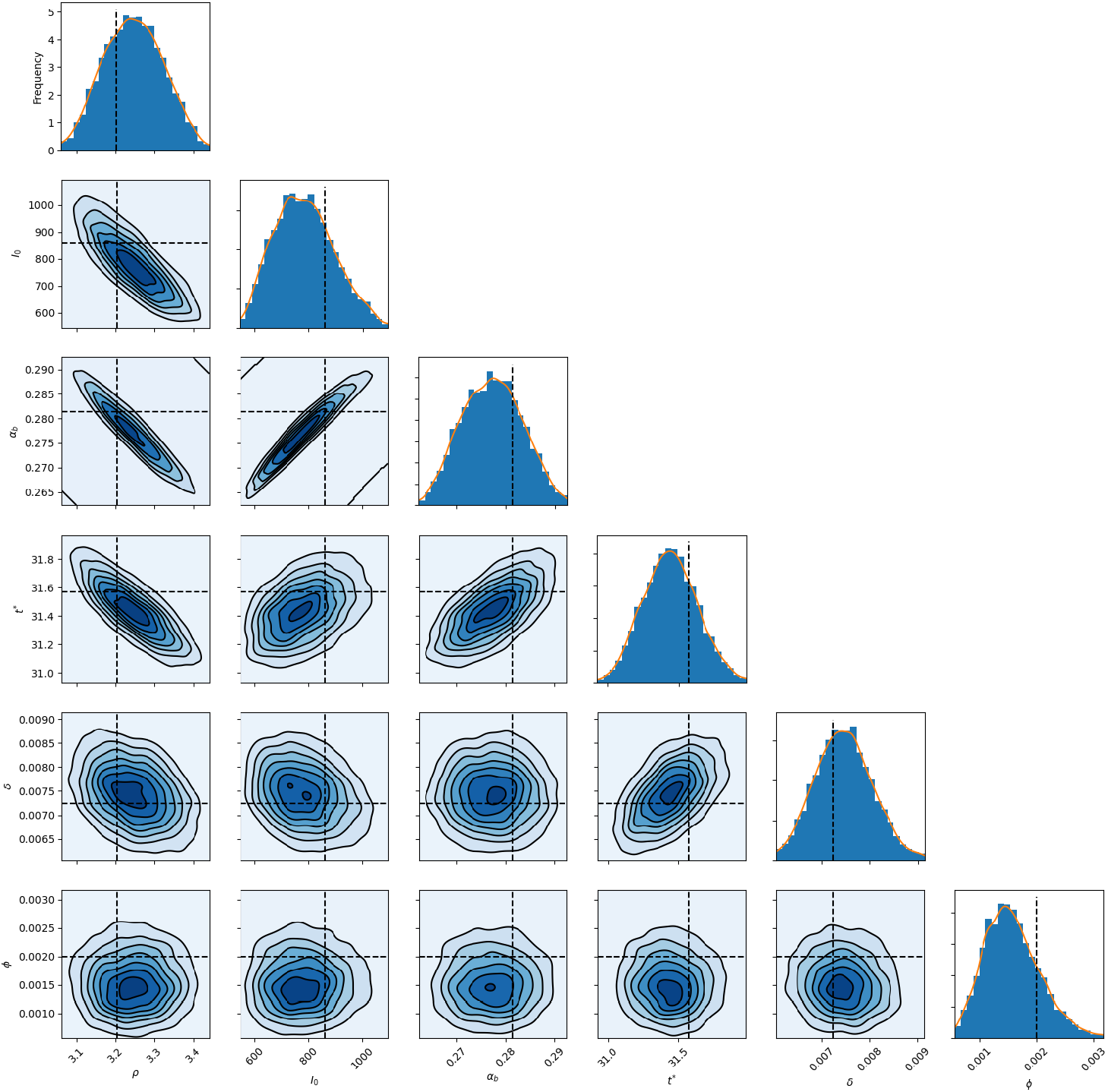
Bayesian inference of synthetic data from Figure 4 using parameters described in §3.3. Uni- and bivariate marginal posterior distributions for the model parameters are shown, with the true, data-generating values shown as black, dotted lines.

**Figure S3:**
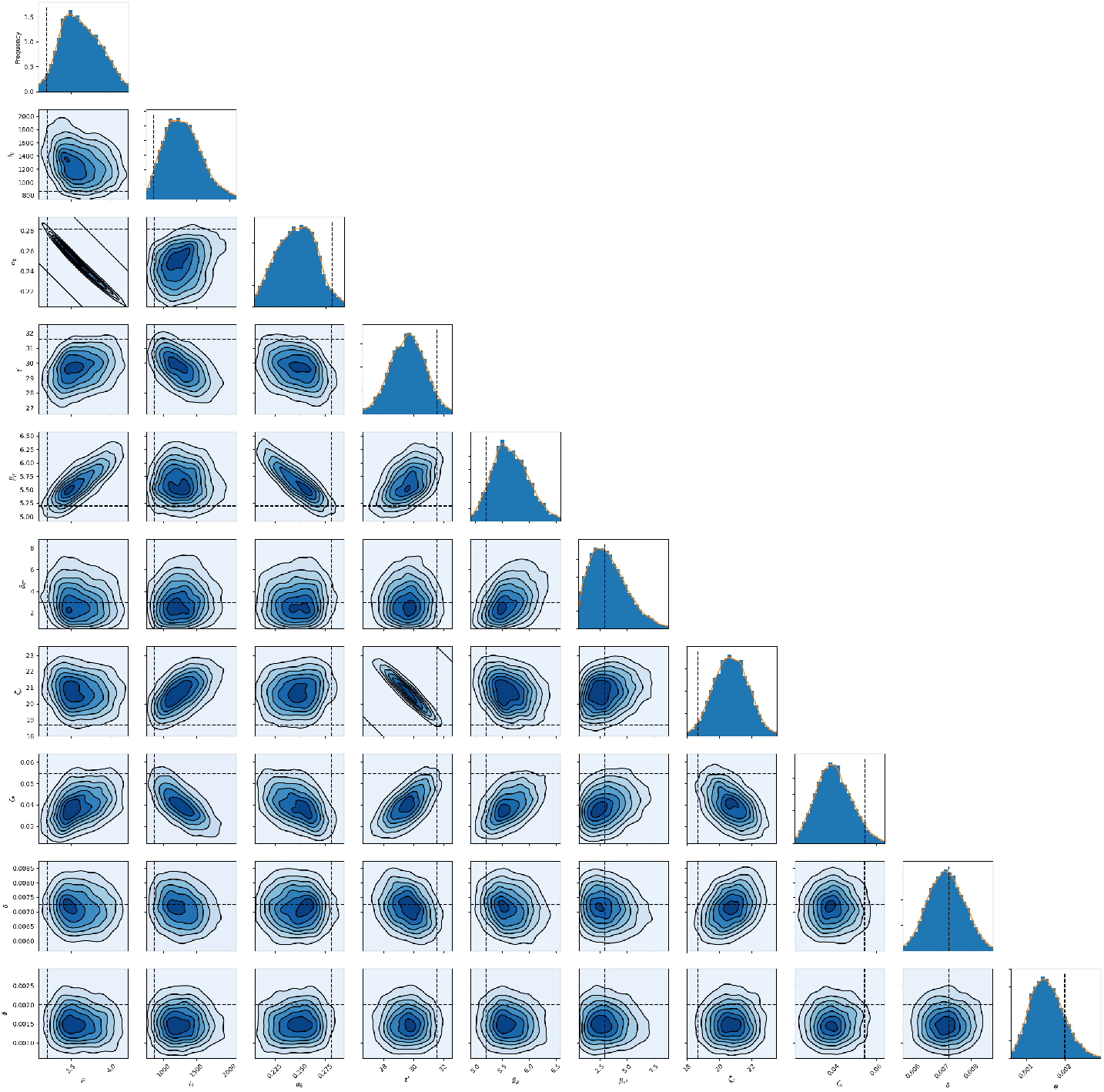
Bayesian inference of synthetic data from Figure 4 using parameters described in §3.4. Uni- and bivariate marginal posterior distributions for the model parameters are shown, with the true, data-generating values shown as black, dotted lines. These results are analogous to those in Fig S2, except here the analysis involved placing priors on—rather than conditioning on—the parameters 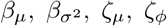 of the infectiousness profile and infection-to-death distribution.

**Figure S4:**
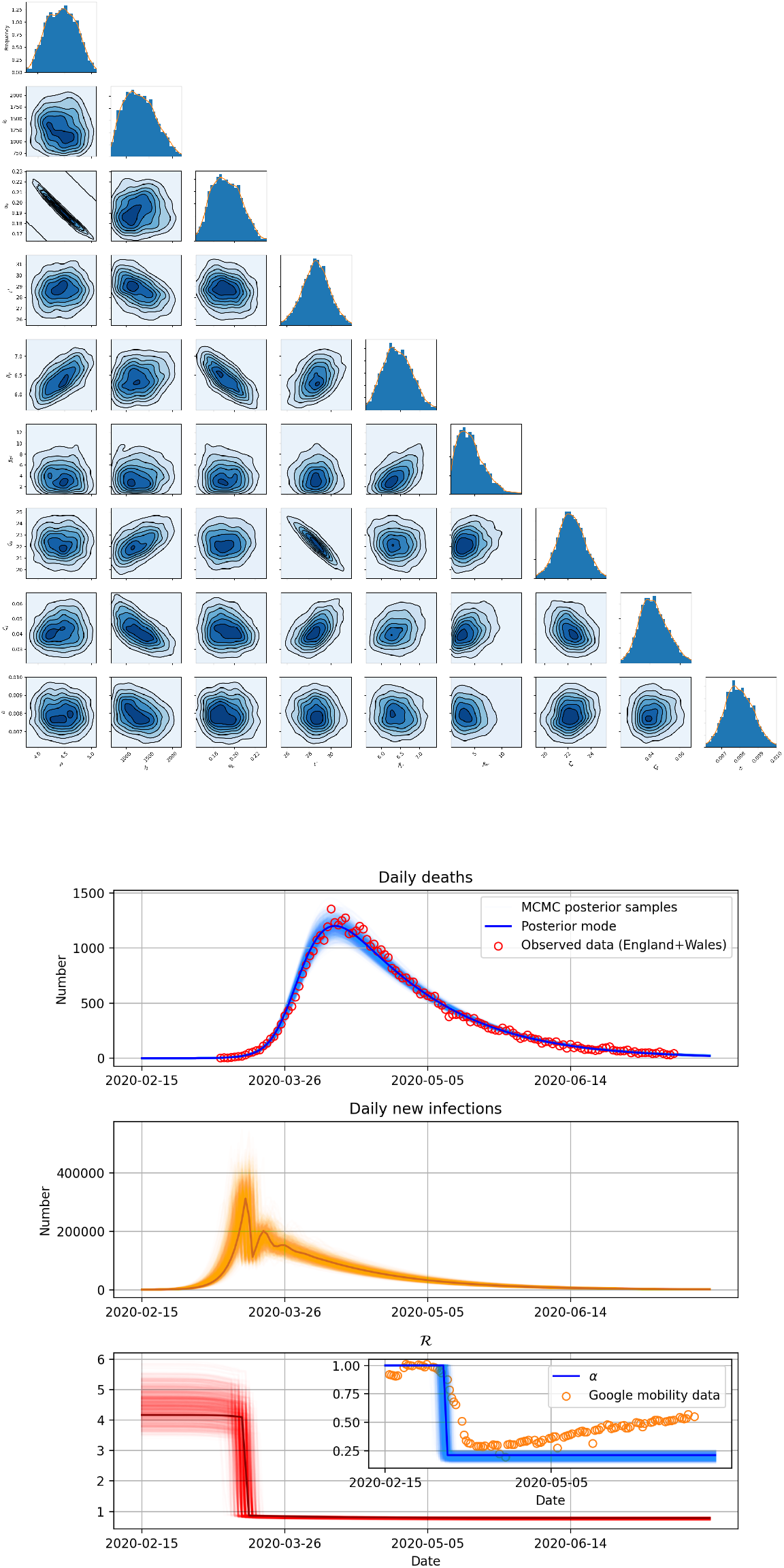
Bayesian inference of real data using the *SI*_*t*_*D* model with uncertainty on 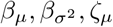 and *ζ*_*ϕ*_, with *ϕ* fixed to 0.01. (Upper panel) Uni- and bivariate marginal posterior distributions for the model parameters. (Lower panel) The interpretation is the same as that of panels (B-D) in Figure 6.

**Figure S5:**
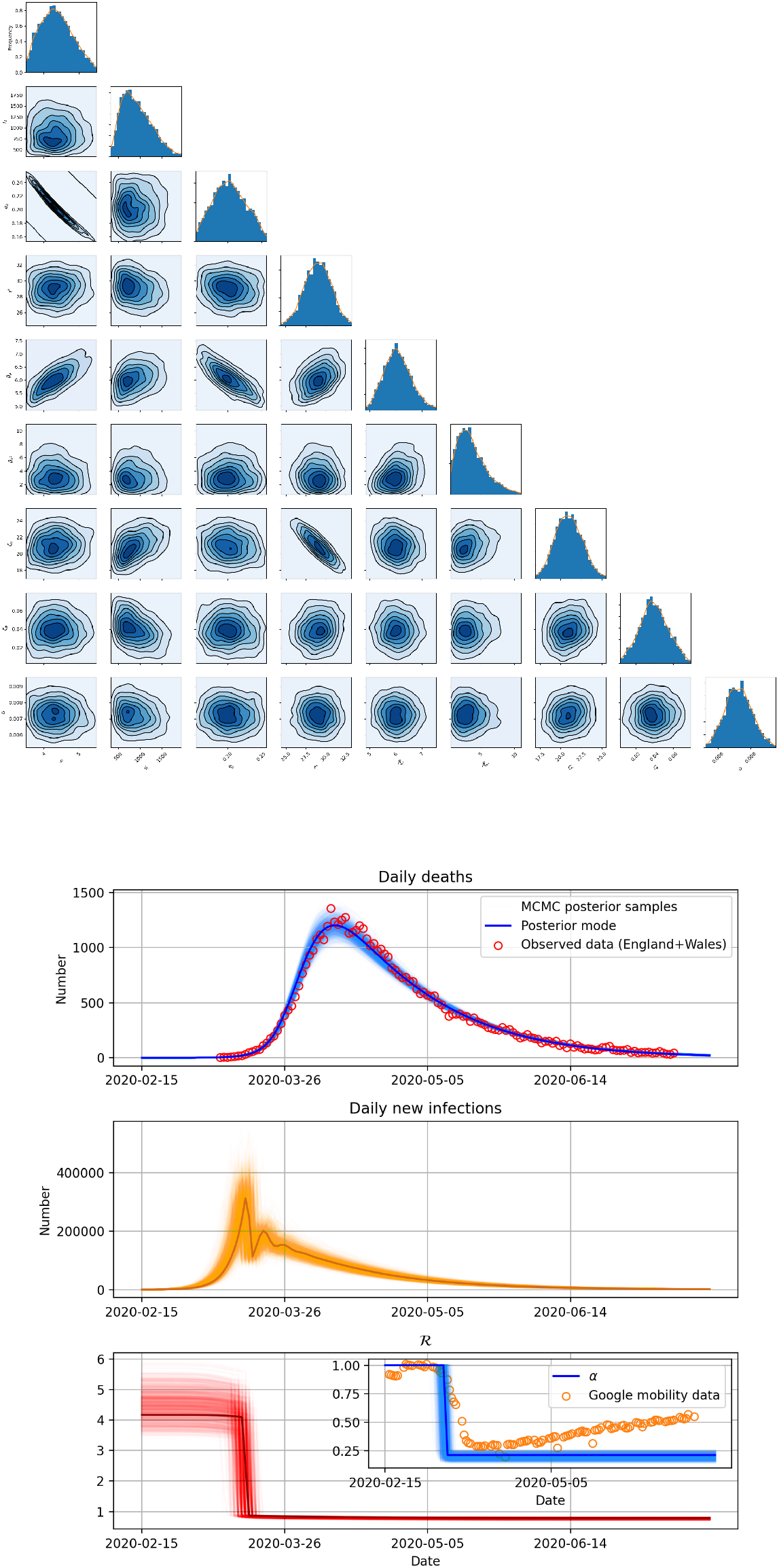
Bayesian inference of real data using the *SI*_*t*_*D* model with 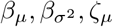 and *ζ*_*ϕ*_, with *ϕ* fixed to 0.1. (Upper panel) Uni- and bivariate marginal posterior distributions for the model parameters. (Lower panel) The interpretation is the same as that of panels (B-D) in Figure 6.

**Figure S6:**
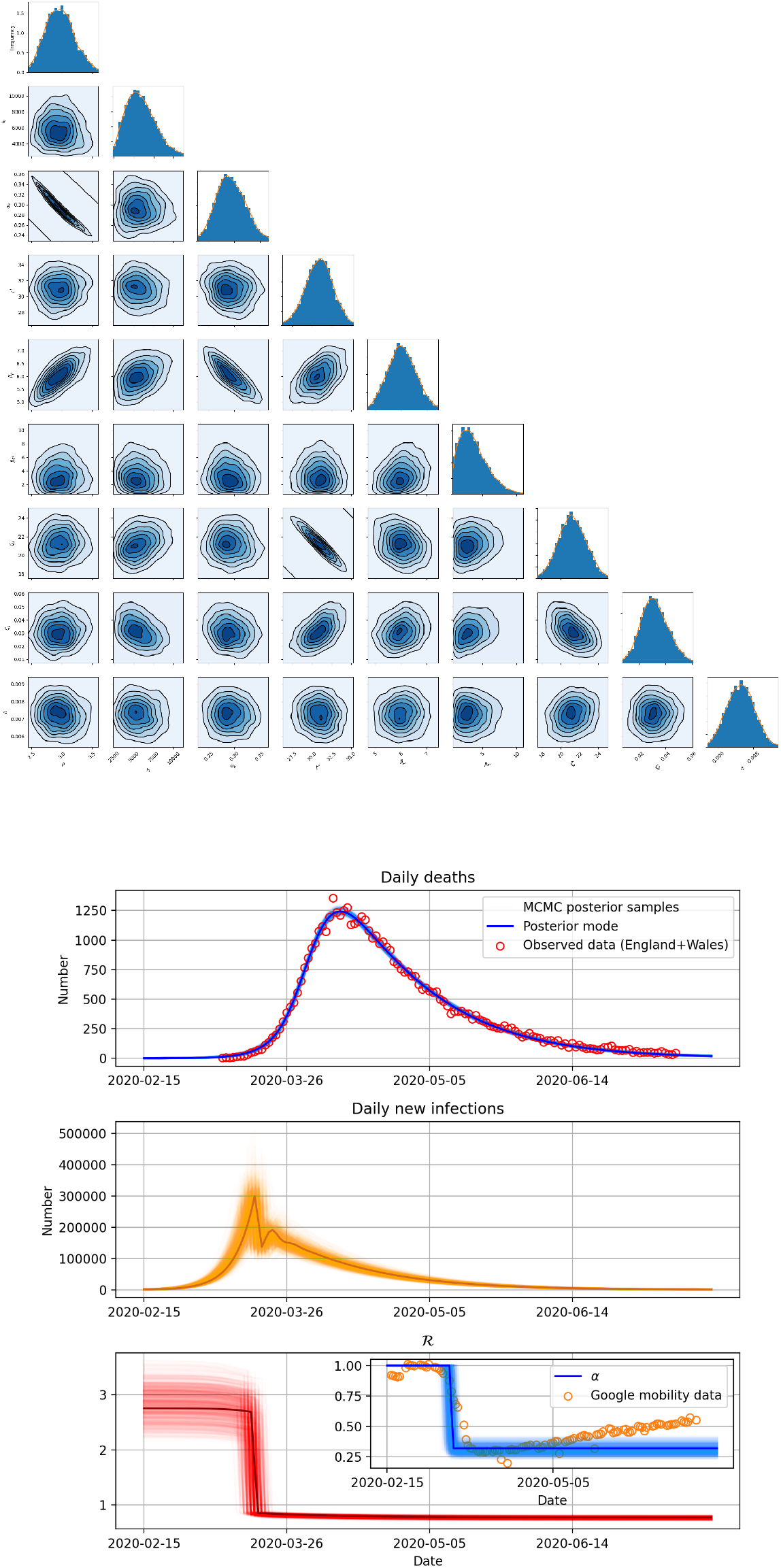
Bayesian inference of real data using the *SI*_*t*_*D* model with uncertainty on 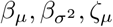 and *ζ*_*ϕ*_, with Gaussian noise *σ* fixed to 60. (Upper panel) Uni- and bivariate marginal posterior distributions for the model parameters. (Lower panel) The interpretation is the same as that of panels (B-D) in Figure 6.

## Notes

### Competing Interest Statement

DGW is now employed by GlaxoSmithKline Plc. There are no other competing interests.

